# Mutational signatures in esophageal squamous cell carcinoma from eight countries of varying incidence

**DOI:** 10.1101/2021.04.29.21255920

**Authors:** Sarah Moody, Sergey Senkin, S M Ashiqul Islam, Jingwei Wang, Dariush Nasrollahzadeh, Ricardo Cortez Cardoso Penha, Stephen Fitzgerald, Erik N Bergstrom, Joshua Atkins, Yudou He, Azhar Khandekar, Karl Smith-Byrne, Christine Carreira, Valerie Gaborieau, Calli Latimer, Emily Thomas, Irina Abnizova, Pauline E Bucciarelli, David Jones, Jon W Teague, Behnoush Abedi-Ardekani, Stefano Serra, Jean-Yves Scoazec, Hiva Saffar, Farid Azmoudeh-Ardelan, Masoud Sotoudeh, Arash Nikmanesh, Michael Eden, Paul Richman, Lia S Campos, Rebecca C Fitzgerald, Luis Felipe Ribeiro, Charles Dzamalala, Blandina Theophil Mmbaga, Tatsuhiro Shibata, Diana Menya, Alisa M Goldstein, Nan Hu, Reza Malekzadeh, Abdolreza Fazel, Valerie McCormack, James McKay, Sandra Perdomo, Ghislaine Scelo, Estelle Chanudet, Laura Humphreys, Ludmil B Alexandrov, Paul Brennan, Michael R Stratton

**Affiliations:** Cancer, Ageing and Somatic Mutation, Wellcome Sanger Institute, Wellcome Genome Campus, Cambridge, CB10 1SA, UK; International Agency for Research on Cancer/ World Health Organization (IARC/WHO), 150 Cours Albert Thomas, Lyon, 69372, France; Moores Cancer Centre, UC San Diego Health, La Jolla, California, 92037, USA; Department of Cellular and Molecular Medicine, University of California, La Jolla, California, 92093; Department of Bioengineering, University of California, La Jolla, California, 92093, USA; Digestive Oncology Research Center, Digestive Diseases Research Institute, Tehran University of Medical Sciences, Shariati Hospital, Tehran, Iran; University Health Network, Toronto, Canada; Department Laboratory Medicine and Pathology, Gustave Roussy, Paris, France; Department of Pathology, Shariati Hospital, Tehran University of Medical Sciences, Tehran, Iran; Liver Transplantation Research Center, Imam Khomeini Hospital Complex, Tehran University of Medical Sciences, Tehran, Iran; Digestive Diseases Research Centre, Digestive Diseases Research Institute, Tehran University of Medical Sciences, Tehran, Iran; Cambridge University Hospitals NHS Foundation Trust, Cambridge Biomedical Campus, Cambridge, CB2 0QQ, UK; Histopathology Department, Hemel Hempstead General Hospital, Hemel Hempstead, HP2 4AD, UK; West Suffolk NHS Foundation Trust, Bury St Edmunds, IP33 2QZ, UK; MRC Cancer Unit, University of Cambridge, Cambridge, CB2 0XZ, UK; Brazilian National Cancer Institute, Brazil; University of Malawi, Malawi; Kilimanjaro Clinical Research Institute, Kilimanjaro Christian Medical Centre & Kilimanjaro Christian Medical University College, Tanzania; Division of Cancer Genomics, National Cancer Centre Research Institute, Tokyo, Japan; Moi University, Kenya; Division of Cancer Epidemiology and Genetics, National Cancer Institute, NIH, USA; Digestive Oncology Research Center, Tehran University of Medical Sciences, Tehran, Iran; Golestan University of Medical Sciences, Iran; Cancer Epidemiology Unit, Department of Medical Sciences, University of Turin, Italy

## Abstract

Esophageal squamous cell carcinoma (ESCC) shows a remarkable variation in incidence which is not fully explained by known lifestyle and environmental risk factors. It has been speculated that an unknown exogenous exposure(s) could be responsible. Here we combine the fields of mutational signature analysis with cancer epidemiology to study 552 ESCC genomes from eight countries with varying incidence rates. The mutational profiles of ESCC were similar across all countries studied. Associations between specific mutational signatures and ESCC risk factors were identified for tobacco, alcohol, opium and germline variants, with modest impacts on mutation burden. We find no evidence of a mutational signature indicative of an exogenous exposure capable of explaining the differences in ESCC incidence. APOBEC associated mutational signatures SBS2 and SBS13 were present in 88% and 91% of cases respectively and accounted for a quarter of the mutation burden on average, indicating that activation of APOBEC is a crucial step in ESCC tumor development.

## Main

Esophageal cancer is the sixth highest cause of cancer deaths worldwide, and esophageal squamous cell carcinoma (ESCC) is the most common subtype^1, 2^. ESCC shows remarkable variation in incidence globally, with the majority of cases occurring in low- and middle-income countries. Regions of particularly high incidence include the Golestan province in Iran, the Shanxi province in China and East Africa^2, 3^. In high income countries the ESCC incidence is generally much lower^2^.

Epidemiological studies have investigated and identified several potential environmental and lifestyle risk factors for ESCC^3–6^. These include tobacco smoking and alcohol consumption, which have been shown to act synergistically to increase the risk of ESCC predominantly in lower risk regions^3, 4, 7^. Polycyclic aromatic hydrocarbon (PAH) exposure from sources other than tobacco smoking, including indoor air pollution and contaminated food and drink, have also been implicated in ESCC risk^3, 4^. Consumption of very hot beverages (including tea drinks and maté in South America), poor diet, poor oral hygiene and opium smoking in Iran are among the other known and speculated ESCC risk factors^3–5^. Despite advances in the understanding of environmental risk factors for ESCC, they are currently considered insufficient to explain the observed international differences in incidence. It has therefore been speculated that an unknown exogenous exposure could be responsible^4^.

The study of mutational processes in human cancers and normal tissues has revealed that both exogenous mutagenic exposures and endogenous processes generate distinctive patterns of mutations, known as mutational signatures^8, 9^. Whilst some of the known mutational signatures have well established biological mechanisms, for others the etiology remains unknown. Similarly, whilst some environmental cancer risk factors have been associated with specific mutational signatures, for others it is currently unknown whether they act directly by generating their own mutational signature, indirectly by influencing the mutation rates of endogenous processes (or a combination of both, as is the case for tobacco smoking), or through non-mutational mechanisms^10, 11^.

If, as hypothesized, there is an unknown specific environmental mutagen driving the large differences in global ESCC incidence, either overall or in a particular setting, then this could be detected by comparing the mutational signatures of ESCC genomes from a variety of locations, spanning the global range of incidence rates. Such a strategy has previously been used to identify a mutational signature present in renal cell carcinomas which implicated an environmental exposure to aristolochic acid in a specific geographic location^12^. Whilst there have been previous studies which have provided a comprehensive overview of the ESCC genome, some including mutational signatures analysis, many of these are limited to whole exome sequencing or cases from high incidence regions only^13–20^. Furthermore, a comprehensive analysis of associations between mutational signatures and ESCC risk factors across multiple regions with varying incidence has not been described previously. Here we investigate further the factors driving global variation in ESCC incidence rates by combining whole genome sequencing and mutational signature analysis with extensive epidemiological questionnaire data, in ESCC from eight countries with varying incidence rates.^4^

## Results

### Data collection

ESCC from high incidence regions (incidence rates ranging from 20/100,000 to 30/100,000, in Iran (Golestan province), Kenya, Tanzania and Malawi and up to 100/100,000 in China (Shanxi province)), and low incidence regions (incidence rates below 6/100,000, Brazil, Japan and UK) were selected for this study^4^. 552 cases were included in the final analysis from the following countries: Brazil (30), China (138), Iran (178), Japan (37), Kenya (68), Malawi (59), Tanzania (35) and United Kingdom (7) (Fig. 1, Methods). Mean sequencing coverage for tumor and germline DNA was 49-fold and 26-fold respectively. Epidemiological questionnaire data was available for all cases on tobacco smoking and alcohol consumption, and in the high incidence countries for hot food and drink consumption, indoor air pollution and opioid usage (Iran only) (Fig. 1).

**Fig. 1:**
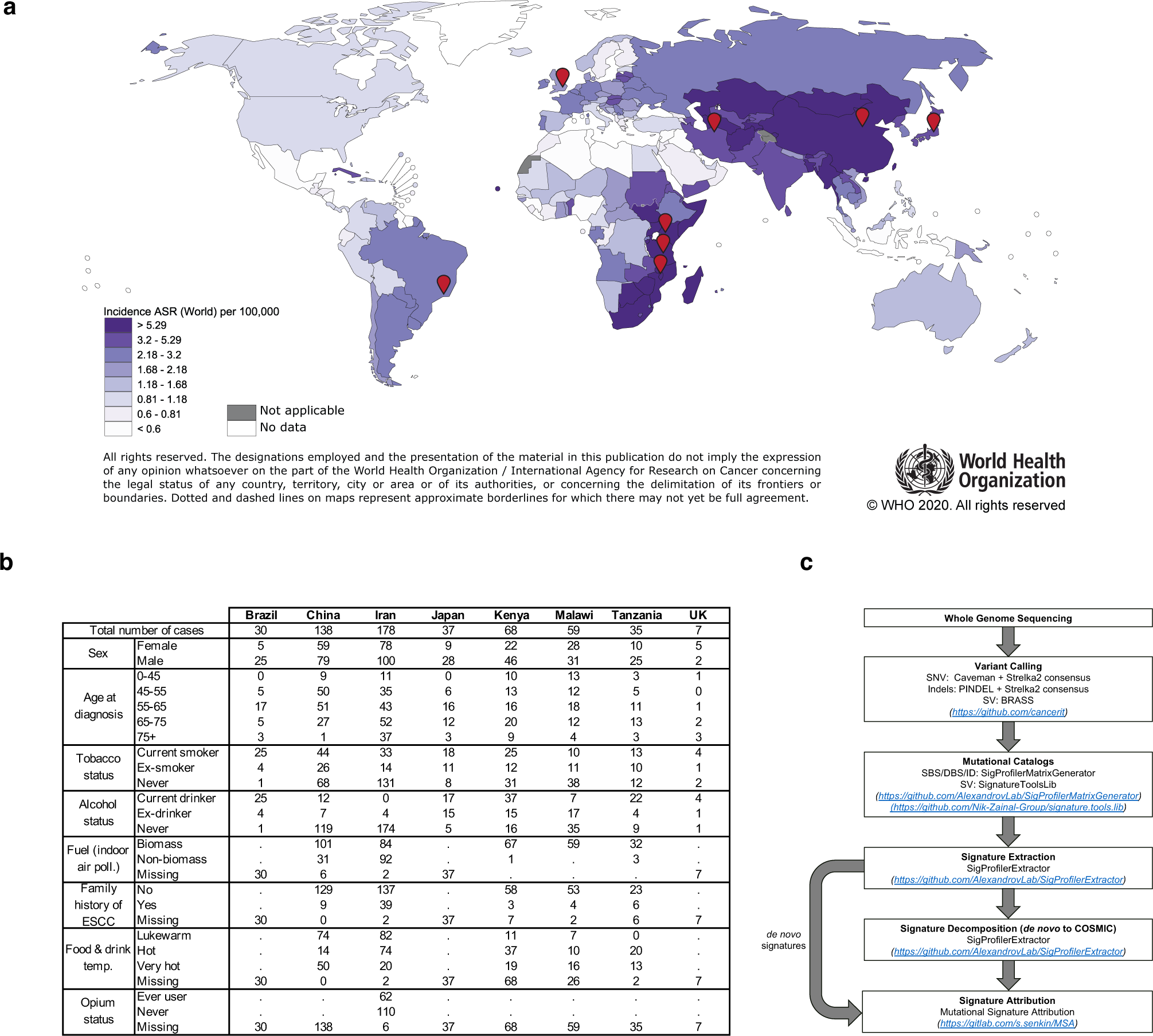
ESCC incidence and data collection. (a) Incidence of esophageal squamous cell carcinoma (sex combined, age standardized rates per 100,000, data from GLOBOCAN 2018). Markers indicate countries included in this study. (b) Summary of ESCC risk factors in the series included in this study (c) Flowchart of methodology used to analyze mutational signatures in this study.

### Mutation burden

Mutation burden ranged from 1191 to 62240 (median 10709) for single base substitutions (SBS) and from 2 to 260 (median 35) for dinucleotide substitutions. Indel mutation burden ranged from 27 to 64358 (median 753). After excluding cases defined as hypermutators (including 26 SBS and 19 indel hypermutators), modest differences in the mutation burdens were found between countries (Extended Data Fig. 1, Supplementary Table 1).

### Mutational signatures

Mutational signature analysis was performed using two different methods, SigProfilerExtractor and HDP (Methods)^21^. No signature indicative of a major unknown exogenous mutagenic exposure accounting for a substantial portion of the mutation burden was found using either method (Fig.2, Extended Data Fig. 2). The SigProfilerExtractor results were used for all further analysis.

**Fig. 2:**
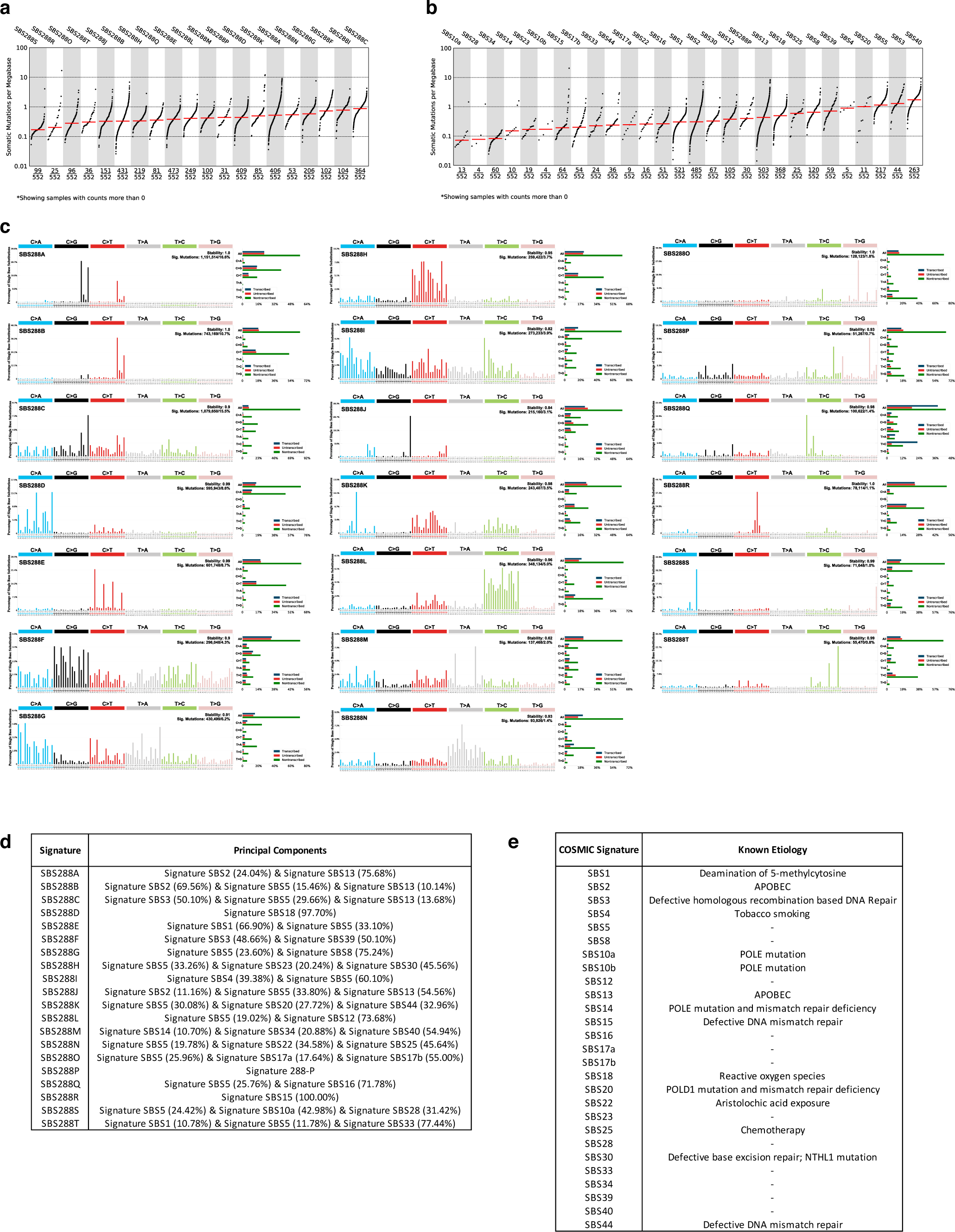
SBS288 mutational signature analysis of ESCC. (a) TMB plot showing the frequency and mutations/mb for each of the extracted *de novo* signatures. (b) TMB showing the frequency and mutations/mb for each COSMIC reference signature identified in the ESCC cohort. (c) Twenty *de novo* signatures extracted from 552 ESCC cases. (d) Mapping of de novo signatures extracted from ESCC decomposed into COSMIC reference signatures. For clarity, components amounting to less than 10% of the signature have been omitted. (e) COSMIC reference signatures identified in the ESCC cohort with known etiologies.

We extracted *de novo* base substitution mutational signatures with SigProfilerExtractor using a mutation classification composed of 288 base substitution types that incorporates the six mutation classes, 3’ and 5’ sequence context of the mutated base and whether the mutation was in non-transcribed DNA, on the transcribed strand of a gene or on the non-transcribed strand of the gene. SigProfilerExtractor extracted twenty *de novo* signatures, which were then collapsed into SBS96 contexts and decomposed into twenty-seven COSMIC reference signatures and one novel signature (which could not be explained by any combination of the existing reference signatures) (Fig. 2, Supplementary Tables 2,6,8,9). Using these decompositions, the mutation burden for each COSMIC signature in each sample was estimated.

Six COSMIC reference signatures were responsible for >80% of the average ESCC mutation burden, SBS1, SBS2, SBS5, SBS13, SBS18 and SBS40. Both SBS2 and SBS13 are thought to be due to activity of the APOBEC family of cytosine deaminases and were present in the majority of cases (88% and 91%) (Fig. 2b), together accounting on average for 25% of the mutation burden^8^. SBS1, associated with spontaneous deamination of 5-methylcytosine, and SBS18, thought to be caused by reactive oxygen species, were present in 94% and 67% of cases and accounted on average for a further 8% and 11% of the mutation burden respectively^8^. The other two signatures, SBS5 and SBS40, are relatively flat signatures which are difficult to reliably attribute, and combined were present in 76% of cases and on average accounted for 38% of the mutation burden. The etiology of SBS5 and SBS40 is unknown, but both have been previously shown to correlate with age^8, 9^. The remaining signatures were present at low levels (<5% average) or only found in rare cases such as those with DNA mismatch repair deficiency. COSMIC signature SBS3, associated with defective homologous recombination-based DNA damage repair, has not been previously found in ESCC, but was found here in 44/552 (8%) cases.

Extraction of dinucleotide signatures identified four *de novo* signatures, which decomposed into four COSMIC reference signatures (DBS2, DBS4, DBS6 and DBS9) and one novel signature (Extended Data Fig. 3, Supplementary Tables 3,6,8,9, Supplementary Results). DBS4, associated with age, and DBS2, associated with both age and exposure to tobacco smoke and other mutagens, were present in 93% and 78% of cases and contributed 34% and 16% on average respectively^8^. DBS6 and DBS9 which both have unknown etiology, were present in 53% and 36% of cases and on average contributed 12% and 12% respectively).

**Fig. 3:**
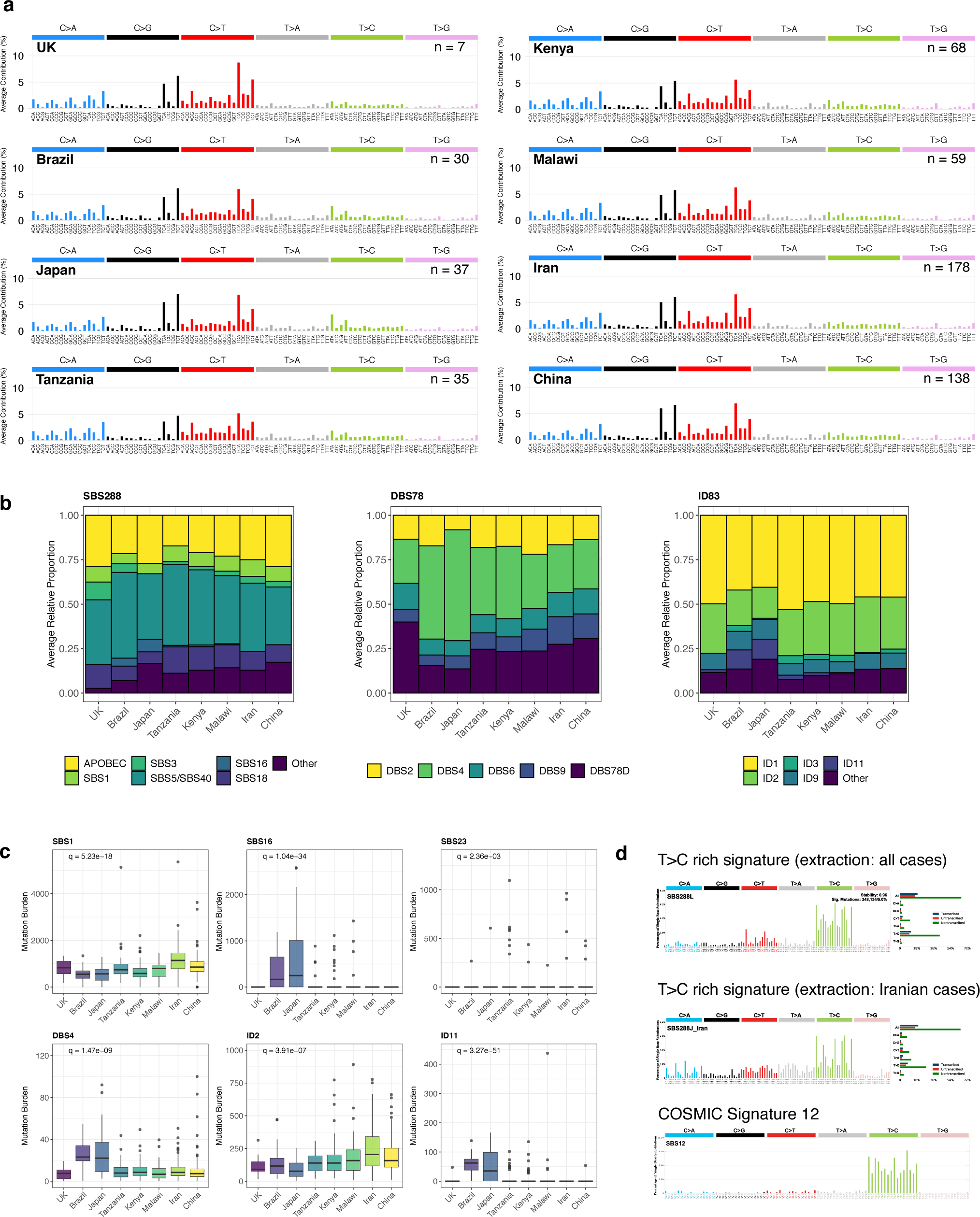
Mutational signatures in ESCC from eight countries. (a) Average SBS96 spectra in ESCC from eight countries are remarkably similar. Countries are ordered by approximate incidence rate in ascending order. (b) Average relative attributions of SBS, DBS and ID COSMIC signatures are broadly similar between all countries. Signatures accounting for less than 5% on average (with the exception of SBS3 and SBS16) are grouped together into the ‘Other’ category. Countries are ordered by approximate incidence rate in ascending order. (c) Mutational signatures showing significant differences in attributed mutation burden between countries. The Kruskal-Wallis test was used to test for global differences. The Bonferroni method was used to adjust p values for multiple hypothesis testing (a total of 53 comparisons). In order to exclude the potential for false positives, significant differences were only declared where the relative attribution, absolute attribution, and absolute attribution following application of confidence intervals were all significant. For clarity, cases with >1000 mutations attributed to ID2 are not displayed. Box and whiskers plots are in the style of Tukey. The line within the box is plotted at the median while the upper and lower ends are indicated 25th and 75th percentiles. Whiskers show 1.5*IQR (interquartile range) and values outside it are shown as individual data points. Countries are ordered by approximate incidence rate in ascending order. (d) Comparison between T>C rich *de novo* signature extracted from all cases, the T>C rich *de novo* signature extracted from Iranian cases and COSMIC signature 12, showing that the T>C component of the Iranian signature is distinct from both the corresponding signature extracted from all cases and COSMIC signature 12.

Extraction of indel mutational signatures identified eight *de novo* signatures, which decomposed into 11 COSMIC reference signatures (Extended Data Fig. 4, Supplementary Tables 4,6,8,9). The dominant signatures were ID1 and ID2, which are attributed to polymerase slippage of the nascent (ID1) or template (ID2) strand. These two signatures were present in 100% and 99% of cases and accounted on average for 47% and 28% of the indel mutation burden respectively. The remaining signatures were present at lower levels (<10%).

**Fig. 4:**
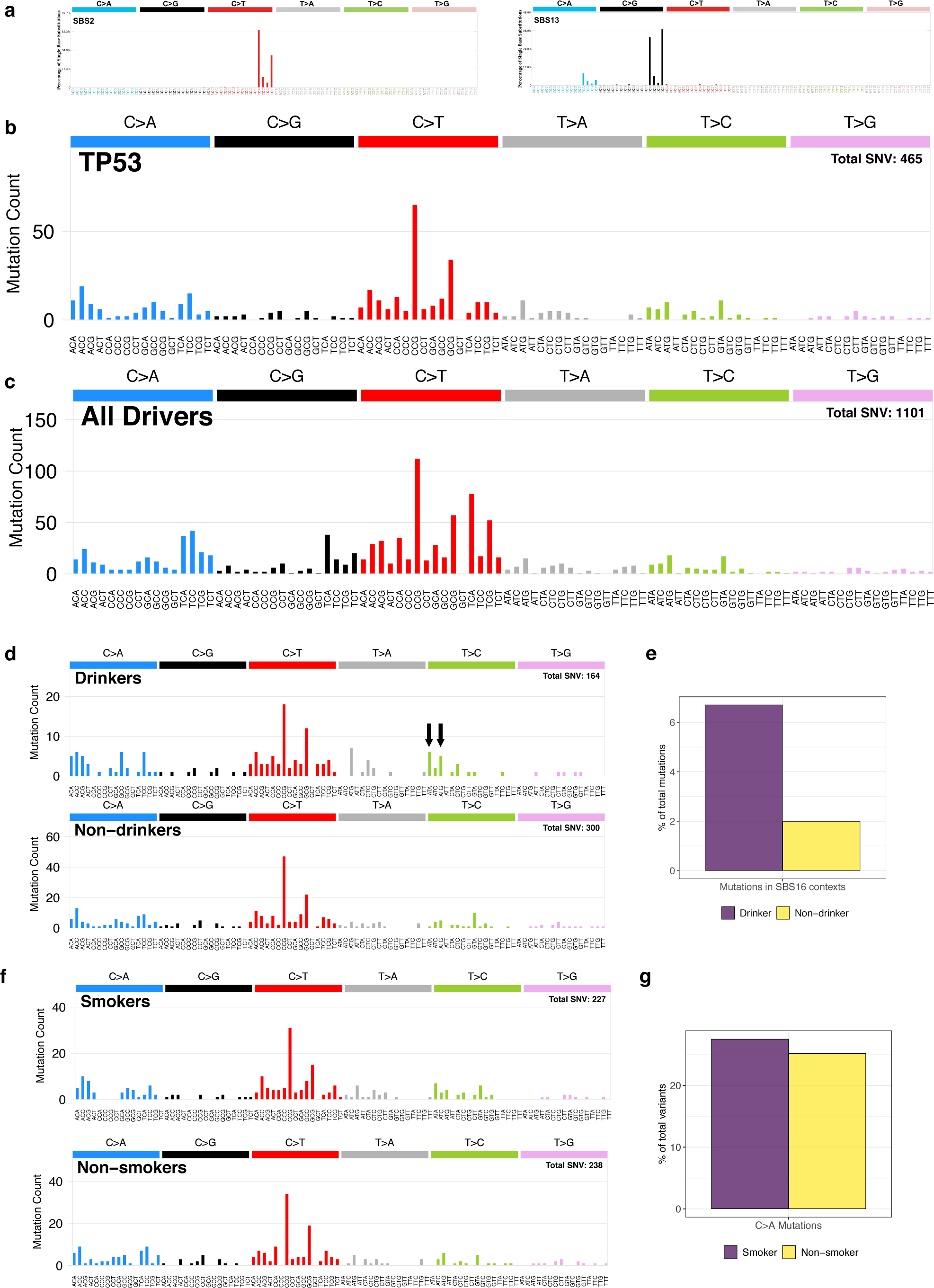
Mutational spectra of ESCC drivers. (a) Spectra of the APOBEC associated COSMIC reference signatures SBS2 and SBS13. (b) SBS96 mutation spectrum of *TP53* mutations in ESCC. A total of 465 SBS variants were identified in 416 cases. The majority of *TP53* mutations are consistent with endogenous mutational processes, with little evidence of APOBEC associated mutational signatures. (c) SBS96 mutation spectrum of all driver mutations in ESCC show some evidence of APOBEC associated mutational signatures. (d) Spectra of *TP53* mutations in drinkers and non-drinkers, showing enrichment of *TP53* mutations in contexts consistent with SBS16 (indicated with arrows). (e) Percentage of *TP53* mutations occurring in SBS16 compatible contexts in drinkers and non-drinkers. (f) Spectra of *TP53* mutations in smokers and non-smokers were similar. g) Percentage of *TP53* mutations occurring in C>A contexts in smokers and non-smokers.

### Mutational signatures in ESCC from regions of varying incidence

Despite the substantial differences observed in ESCC incidence rates, the average mutational spectra of ESCC from different regions, including those from lower incidence regions, were very similar (Fig.3a/b, Extended Data Fig. 5). For SBS288 extractions no significant differences in the attributed mutation burden were seen in five of the dominant signatures (SBS2, SBS5, SBS13, SBS18 and SBS40), whilst significant differences were seen in SBS1 (q=5.23*10^−18^, Supplementary Tables 9, 10). Significant differences were also identified in SBS16 and SBS23 (q=1.04*10^-34^ and q=2.36*10^-3^, Fig. 3c). SBS16 has previously been associated with alcohol consumption in ESCC, and SBS16 mutation burden was significantly higher in the lower incidence countries Brazil and Japan when compared to all other regions (p=5.06*10^-15^ and p=1.68*10^-23^ respectively, Extended Data Fig. 6)^18, 19^. The etiology of SBS23 is unknown, and was only present in a small subset of cases with a modest mutation burden (19/552 (3%), median = 475, Fig. 3c). For DBS signatures significant differences were only observed in DBS4 (q=1.47*10^-9^), which was enriched in Brazil and Japan when compared to all other regions (p=3.67*10^-8^ and p=4.92*10^-7^ respectively, (Extended Data Fig.6, Supplementary Tables 9, 10). Significant differences were found in ID2 and ID11 (q=3.91*10^-7^ and q=3.27*10^-51^, Fig. 3c, Supplementary Tables 9, 10). The observed differences in both ID2 and SBS1 are likely explained by age, as both are significantly enriched in Iran compared to most other regions (p=2.55*10^-7^ and p=2.51*10^-17^ respectively, Extended Data Fig.6), which correspondingly has a modestly increased average age at diagnosis compared to most other regions. ID11 was largely restricted to Brazil and Japan, and was significantly enriched in these regions (p=6.18*10^-30^ and p=4.35*10^-23^ respectively, Extended Data Fig. 6).

**Fig. 5:**
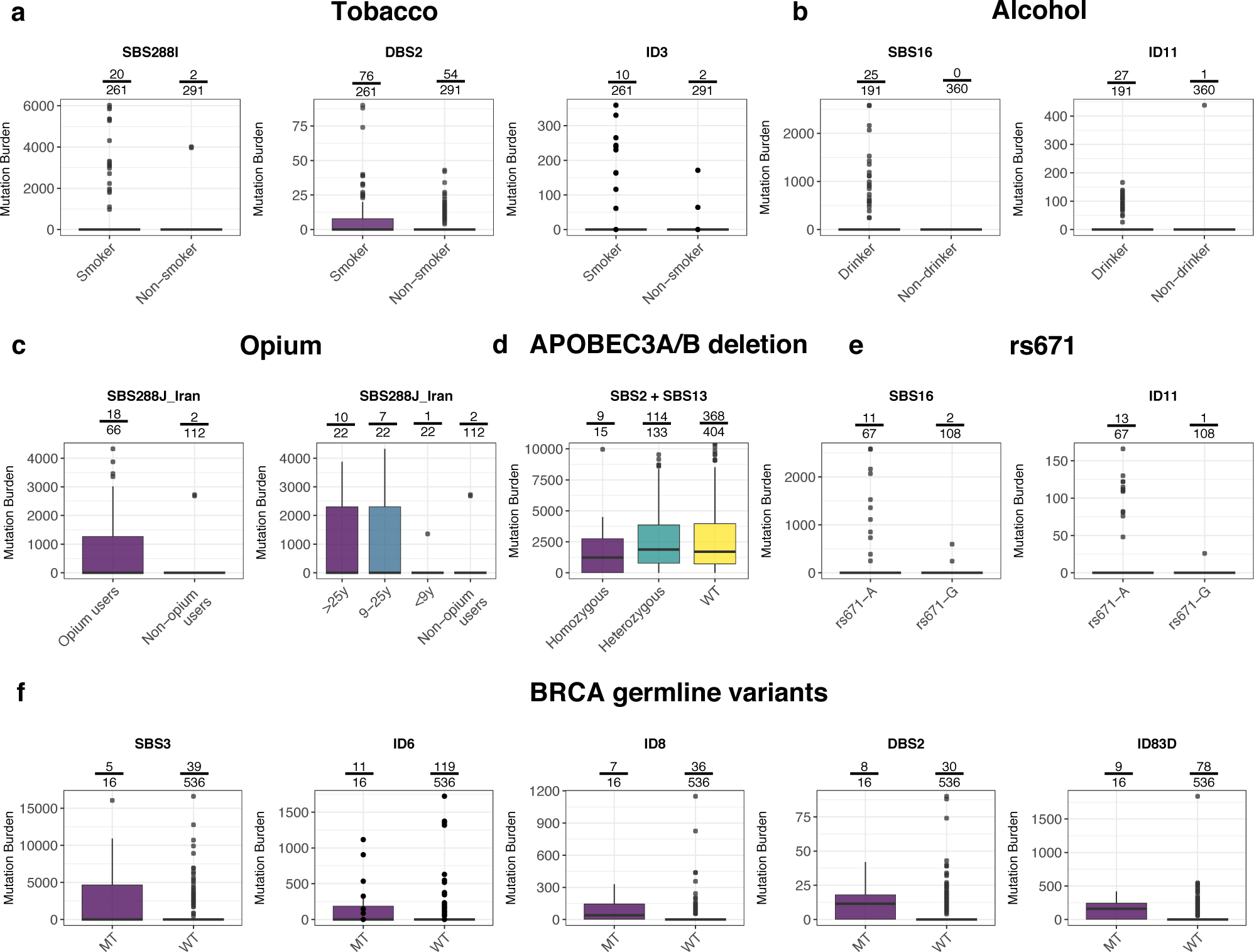
Associations between ESCC risk factors and mutational signatures. Number of mutations attributed to mutational signatures with respect to (a) tobacco smoking status for signatures SBS288I, DBS2 and ID3; (b) alcohol drinking status for signatures SBS16 and ID11; (c) opium ever-use status and opium duration of use for signature SBS288J; (d) APOBEC3A/B germline deletion genotype for merged APOBEC signatures SBS2 and SBS13. For clarity, cases with >10,000 mutations attributed to APOBEC are not displayed; (e) rs671 germline variant genotype for signature SBS16; (f) putative deleterious germline BRCA1/2 variants genotype for signatures SBS3, DBS2, ID6, ID8 and ID83D. Box and whiskers plots are in the style of Tukey. The line within the box is plotted at the median while the upper and lower ends are indicated 25th and 75th percentiles. Whiskers show 1.5*IQR (interquartile range) and values outside it are shown as individual data points. Labels above columns show the number of cases with >0 mutations attributed vs the total number of cases in the column.

In order to explore the possibility of a mutational signature present predominantly in the high incidence regions that accounted for a relatively small mutation burden, region specific SBS288 signature extractions were performed on three series with >100 cases: from China, East Africa (Kenya, Malawi and Tanzania) and Iran. The extracted signatures from the China and East Africa extractions were similar to those extracted from the full cohort (Extended Data Fig. 7). In the regional extraction results from Iran a difference was found in a T>C rich signature (SBS288J_Iran) compared to the equivalent signature in the complete cohort (Fig. 3d). Both *de novo* signatures decomposed into COSMIC reference signatures SBS1, SBS5 and SBS12, with the regional signature containing an additional component, SBS25. This signature has only been found previously in Hodgkin lymphoma cell lines derived from patients previously treated with chemotherapy, making it unlikely to be present in this context^8^. A more likely scenario is that the regional signature contains a novel component. This signature was present in 47/178 (26%) of Iranian ESCC, and when present contributed between 10-40% (average 17%) of the total mutation burden.

### Mutation Spectra of Driver Mutations

Genes under positive selection were identified using an approach based on the observed to expected ratios of non-synonymous and synonymous mutations (dN/dS)^22^. In total 38 genes were identified (Supplementary Table 11), including known ESCC drivers such as *TP53*, *CDKN2A*, *PIK3CA*, *NFE2L2* and *NOTCH1*. Genes under selection, known cancer genes and ESCC drivers reported elsewhere were then screened for driver mutations (Methods)^13, 17, 18, 22^. In total 1520 driver mutations were identified in 129 genes (Supplementary Table 12).

The most frequently mutated cancer gene in ESCC was *TP53* (500/552, 91% cases), as previously reported^13, 17, 18^. Of these 465 were SBS variants which were used to construct a *TP53* SBS96 mutation spectrum, which showed little evidence of APOBEC mutagenesis (COSMIC signatures SBS2 and SBS13) (Fig. 4b). By comparison, examining the spectrum constructed from all SBS ESCC driver mutations suggests that some drivers arise as a result of APOBEC associated mutational signatures (Fig. 4c). Examining the *TP53* mutation spectrum in alcohol drinkers revealed an enrichment of mutations with the characteristic profile of SBS16 compared to the spectrum in non-drinkers (Fig. 4d-e, p= 0.017), suggesting that alcohol may contribute to mutation of this cancer gene and thus its contribution to cancer development. Despite previous literature demonstrating enrichment of C>A mutations in *TP53,* no difference was observed here between smokers and non-smokers, although this might be confounded by sources of C>A mutations other than tobacco smoke (Fig. 4f-g).

### Clonality of APOBEC mutations

In order to address the question of whether APOBEC mutations are clonal, and therefore occur relatively early in the development of ESCC, the variant allele frequency (VAF) was examined in C>G and C>T mutations at TCN (T base at the 5’ position and any base at the 3’ position) contexts (Methods). A lower VAF than other mutations would indicate that APOBEC mutations are more commonly subclonal. However, no significant differences in the VAF profile of C>G and C>T mutations at TCN contexts was found either in all cohorts combined or in individual countries (Extended Data Fig. 8, Supplementary Table 13).

### Associations with risk factors

Regression analysis was performed to identify associations between the mutation burdens attributed to the mutational signatures present in each case and epidemiological risk factors. Regressions were carried out using attributions to both *de novo* and COSMIC signatures. Where possible the results are presented here using COSMIC attributions (Supplementary Tables 14-16).

No association was observed between any risk factors and the APOBEC signatures. However, associations between several other signatures and ESCC risk factors were found, albeit all with modest gains in mutation burden. Tobacco smoking was associated with COSMIC signatures DBS2 (p=3.2*10^-5^) and ID3 (p=0.017) as previously reported (Fig. 5a)^8^. No COSMIC SBS signature was associated with smoking, however *de novo* signature SBS288I (of which SBS4 is a component) did show an association (p=9.8*10^-6^, Fig. 5b). This is likely due to the low mutation burden attributed to SBS4, whereas the *de novo* signature SBS288I is a composite of multiple mutational signatures affected by smoking either directly or indirectly. Alcohol consumption was associated with COSMIC signatures SBS16 and ID11 (p=1.8*10^-3^, p=0.036), which were exclusively attributed in drinkers, with the exception of one case with ID11 (Fig. 5b). The association between alcohol consumption and SBS16 has been previously reported in ESCC from China and Japan^18, 19^, but association with ID11 has not been previously reported. These signatures also strongly associated with living in areas of lower ESCC incidence (Brazil, Japan and United Kingdom; p=1.8*10^-7^, p=1.4*10^-8^) where alcohol has been reported as an important risk factor for ESCC. These signatures were also found in East African cases. However, statistical power was limited to disentangle the effects of tobacco and alcohol consumption in these cases. No association between family history of ESCC, the use of biomass fuels, (an indicator for indoor air pollution) or hot food and drinks was observed with COSMIC or *de novo* signatures.

Data on opium use was available for the Iranian cohort, and *de novo* signature SBS288J_Iran was associated with opium exposure (p=9.5*10^-5^), as well as the duration of opium use (trend p=3.6*10^-7^, Fig. 5c). This is the T>C enriched signature which showed differences in its composition compared to the corresponding signature from all ESCC (Fig. 2). This signature is speculated to contain a novel component, with the observed association suggesting that this could potentially be driven by opium exposure. However, as it was not possible to extract this component independently of other COSMIC signatures, it is not possible to confirm this.

### Associations with germline variants

Although little is known about germline susceptibility to ESCC, germline variants previously implicated in ESCC risk were investigated to identify any associations with mutational signatures. Analysis was performed on both *de novo* and COSMIC reference signatures, and where possible the results are presented here using COSMIC attributions (Supplementary Tables 14,15,17).

A germline 30kb deletion in the APOBEC3 region has been previously implicated both in cancer risk and in influencing APOBEC mutagenesis, and was detected in 148/552 (27%) cases, of which fifteen were homozygous deletions^23–25^. The deletion was associated with decreased total APOBEC mutation burden (COSMIC signatures SBS2 and SBS13) (p=0.007, p=1.0*10^-4^, Fig. 5d), in contrast to previous studies which have observed increased overall APOBEC mutagenesis in carriers^23, 24, 26^. In ESCC from China and Japan, the frequency of rs12628403, which acts as a proxy for the deletion was lower than expected in the general East Asian population (0.28 vs 0.34)^27^, supporting a potential protective role in ESCC, although this variant appeared very rare or understudied in other populations.

*ALDH2* encodes aldehyde dehydrogenase, a key enzyme involved in the metabolism of alcohol which catalyzes the conversion of acetaldehyde to acetic acid. A germline variant in *ALDH2*, rs671(G>A) has been observed almost exclusively in Chinese and Japanese populations and both heterozygote and homozygous carriers have a reduced ability to metabolize acetaldehyde and display toxic reactions following alcohol consumption. Heterozygous rs671-A individuals have been observed to have a substantially greater risk of ESCC if they consume alcohol, likely due to the higher levels of acetaldehyde to which they are exposed^28, 29^. Frequency of the rs671-A allele was enriched in Japanese ESCC, with the genotype distributions skewed towards heterozygotes (GG 8 GA 29 AA 0, pHWE=8*10^-4^), whereas the genotype distributions respected that expected by Hardy-Weinberg equilibrium in the Chinese cohort (GG 100 GA 34 AA 4, pHWE=0.6). Attribution of COSMIC signature SBS16 was significantly increased in ESCC cases with rs671-A as previously reported (p=0.0075)^18, 19^, and COSMIC signature ID11 signature was also significantly increased (p=0.0037, Fig. 5e). Both these associations appear largely driven by patients from Japan, which is likely due to the fact that 86% of cases from Japan reported drinking alcohol at least once per week compared to only 14% of cases from China (Fig.1), consistent with previous reports that alcohol consumption is not as relevant in the Chinese population included in this study^16, 30^.

Finally, germline BRCA2 mutations have been reported to be associated with ESCC in high risk populations from both Iran and China^31, 32^. Putative deleterious germline *BRCA1/2* variants were identified in 16/552 (3%) cases (Supplementary Table 18). Cases with putative deleterious variants were associated with increased attribution of COSMIC signatures SBS3 (p=0.0027), ID6 (p=3.4*10^-6^) and ID8 (p=6.4*10^-7^), signatures which have been previously associated with such mutations in these genes (Fig. 5f)^8^. Interestingly, deleterious germline BRCA variants were also associated with the tobacco-related signatures DBS2 (p=6.2*10^-5^) and ID3 containing *de novo* signature ID83D (p=3.5*10^-5^, Fig. 5f). These results were consistent in cases with somatic BRCA1/2 drivers (n=12), and in both groups combined (Supplementary Table 17).

## Discussion

Despite the profound global differences in ESCC incidence being suggestive of an underlying exogenous exposure, mutational signature analysis of cases from regions of varying incidence did not identify a plausible known or novel mutational process that could be responsible for these differences either overall or in any specific setting. Instead the results show that the overall mutational profile of ESCC is extremely consistent, only deviating in rare cases which are most often due to defects in DNA mismatch repair.

Whilst mutational signatures associated with APOBEC have been identified previously in ESCC, they were found at higher frequency compared to previous studies^15^. A limitation of this study was the difficulty in acquiring large numbers of cases originating from low incidence regions. However, the results presented here are consistent with whole exome sequencing data from other low incidence regions, which also show consistent evidence of APOBEC associated mutational signatures^17, 33^. The near universal presence of APOBEC signatures suggests that activation of APOBEC mutagenesis is an important, and potentially mandatory, step in the genesis of most ESCC. However, when it occurs is unclear. Neoplastic clones of microscopically normal cells with driver mutations in cancer genes such as *NOTCH1*, *NOTCH2* and *TP53* are common in normal esophagus^34, 35^. These normal cell clones have very few rearrangements and copy number changes. Given the similarity in the repertoire of mutated cancer genes between these normal clones and ESCC it is likely that most ESCC originate from these normal clones. Since APOBEC mutational signatures are either absent or only present at low levels in normal clones with driver mutations, it would seem that APOBEC mutagenesis occurs after their formation and expansion and thus may contribute to the rare event of progression of a neoplastic normal clone to a more advanced neoplasm with a more disordered genome. This speculation is supported by the fact that APOBEC signatures are found in dysplastic esophagus^34–36^. However, it is important to note that studies of normal esophagus have been carried out only in UK and Japanese series, and it is currently unknown if these findings are consistent in high incidence regions^34, 35^. The indication that some APOBEC mutations are clonal in ESCC further supports the hypothesis that APOBEC mutagenesis is active in the early stages of ESCC development. The apparent absence of APOBEC mutational signatures in the spectrum of *TP53* mutations confirms that inactivation of *TP53* occurs prior to the onset of APOBEC mutagenesis (albeit this pattern may be skewed by selection). Indeed, it is conceivable that *TP53* mutation is required to enable a permissive environment for the accumulation of APOBEC mutations, and/or for their potential consequences, notably the generation of genomic instability. Additional studies on ESCC tumor evolution will be required to confirm this hypothesis and to further investigate the timing of APOBEC mutations.

We found several associations involving non-APOBEC mutational signatures and ESCC risk factors. Although several of these associations have been reported previously, we did identify novel associations between alcohol consumption and DBS and indel mutational signatures, as well as associations with opium. Opium consumption was recently classified as carcinogenic to humans (Group 1), with limited evidence in esophageal cancers^37^. Our results support a role for opium consumption in ESCC, suggesting that there may be a novel signature associated with its use, although we were unable to fully distinguish it from other signatures with overlapping contexts. It is likely that *in vitro* studies will be needed to clearly establish whether opium consumption generates a mutational signature. Overall, the identified associations had only modest impacts on the total mutation burdens of ESCC. For several other ESCC risk factors no association was found with any mutational signature, with the caveat that the unavailability of certain exposure variables in low incidence regions may have impacted the power to detect associations. The results therefore suggest that some risk factors for ESCC may act by mechanisms other than direct mutagenesis. The idea that known carcinogens can act as non-mutagenic “tumor-promoting” agents is supported by recent studies in mouse models which showed that many do not generate mutational signatures or increase mutation burden^11, 38^. One possibility is that, by causing tissue damage and subsequent wound healing responses, ESCC risk factors increase the number and/or size of normal cell clones with driver mutations and thus increase the risk of a cancer developing from such a clone. Alternatively, ESCC risk factors might increase the frequency of APOBEC machinery activation. Some support for this notion comes from the study of normal esophagus in a Japanese cohort which found that while rare overall, an APOBEC-like signature was enriched in individuals with a history of heavy drinking or smoking^35^.

We also identified several associations between germline variants and mutational signatures. For germline deletions in the APOBEC3 region, previous studies have reported that the deletion was associated with a reduced APOBEC mutation load in cancer types with low APOBEC activity but an increased load in cancer types with high APOBEC activity^26^. The factors driving the apparent discrepancy between our results and previous studies are unclear, but suggest the impact of this deletion is complex and context dependent, and in need of further investigation. Our results confirm the importance of the rs671 and alcohol consumption as being of particular importance in Japan, but not in the Chinese population included in this study^16, 30^. Lastly, our results identify for the first time the presence of mutational signatures associated with defective homologous recombination-based DNA damage repair in ESCC, and establish a role for both germline and somatic *BRCA1/2* variants, suggesting potential therapeutic options for this small subset of cases.

This study, together with the previous finding that some human carcinogens do not generate mutational signatures, highlight a limitation of mutational signature analysis in detecting environmental exposures, with the absence of a distinctive mutational signature being insufficient to rule out the presence of an environmental exposure^11^. It is unlikely that we have failed to detect a SBS signature on the basis of sensitivity, as we estimate that a non-flat signature contributing more than 0.8% of all somatic mutations in the cohort would have been detected by our global analysis. For flat signatures this increases to 3.9% of all mutations, as these are harder to separate and require more somatic mutations in order to be extracted^8^. Importantly, mutational signatures which closely resemble another signature are more difficult to detect and extract (such as the potential opium signature in this study), as the current approach allows the separation of *de novo* mutational signatures only when their cosine similarity is below 0.95^21^. Although we did not find any evidence of a mutational signature capable of explaining the observed differences in incidence rates, we have revealed novel insights into the mechanisms of ESCC risk factors and identified multiple avenues for future studies. We have speculated here that activation of APOBEC mutagenesis appears to be a mandatory step in the oncogenesis of most ESCC, and further study of the factors driving APOBEC activation, as well as in depth studies of ESCC tumor evolution, are likely to yield additional insights.

## Supporting information

Supplementary Results

## Online Methods

### Recruitment of cases

The International Agency for Research on Cancer (IARC/WHO) coordinated case recruitment through an international network of collaborators including Digestive Diseases Research Institute/Tehran University of Medical Sciences, Gonbad, Iran; Shanxi Cancer Hospital, Shanxi, China/ National Cancer Institute, Bethesda, USA; cases within the ESCCAPE case-control studies at Moi Teaching and Referral Hospital/Moi University, Eldoret, Kenya, Queen Elizabeth Central Hospital/University of Malawi, Blantyre, Malawi and Kilimanjaro Clinical Research Institute/Kilimanjaro Christian Medical Centre, Moshi, Tanzania; Brazilian National Cancer Institute, Rio de Janeiro, Brazil and Addenbrooke’s Hospital/MRC Cancer Unit University of Cambridge, Cambridge, United Kingdom (Supplementary Table 19). The inclusion criteria for patients were >=18 years of age, confirmed diagnosis of primary squamous cell carcinoma of the esophagus (ESCC) and no prior treatment. Patients were excluded if they had any condition that could interfere with their ability to provide informed consent or if there were no means of obtaining adequate tissues as per the protocol requirements. Ethical approvals were first obtained from each Local Research Ethics Committee and Federal Ethics Committee when applicable, as well as from the IARC Ethics Committee.

### Informed consent, biosamples and data collection

Dedicated Standard Operating Procedures, following guidelines from the International Cancer Genome Consortium, were designed by IARC/WHO to select adequate ongoing/retrospective case series with complete biological samples and exposure information (Supplementary Table 19). In brief, for all case series included, anthropometric measures were taken, together with relevant information regarding medical and familial history. Blood samples were drawn, and a 30-min questionnaire was administered by a trained interviewer to collect complementary lifestyle and environmental information. Blood collection in EDTA tubes consisted in 3 to 10ml preserved as whole blood or immediately processed into buffy-coat, plasma and red blood cells, and stored at −80 °C. Tumor and matched normal tissues were collected before any treatment and while minimizing routine care disruption. Unless esophagectomy was first line of treatment, mucosal biopsies were collected through upper GI endoscopy. Tumor and matched normal tissues were snap-frozen in liquid nitrogen or promptly preserved in RNAlater® (RNAprotect TissueTubes, QIAGEN). Laboratory Information Management Systems at local recruiting centers, IARC/WHO Biobank and The Wellcome Sanger Institute allowed real-time tracking of biosamples. All patient related data as well as clinical, demographical, lifestyle, pathological and outcome data were pseudonymized locally through the use a dedicated alpha-numerical identifier system before being transferred to IARC/WHO central database. IARC/WHO harmonized all retrospective data based on pre-approved detailed questionnaire(s) and associated data dictionaries (Supplementary Table 20).

### Expert pathology review

Original diagnostic pathology departments provided diagnostic histological details of contributing cases through standard abstract forms, together with a representative hematoxylin-eosin (H&E) stained slide of formalin-fixed paraffin embedded (FFPE) tumor tissues whenever possible. Cases from Japan (37) and the UK (7) completed pathology evaluation locally under the same criteria for tumor eligibility described below. IARC/WHO centralized the entire pathology workflow, and coordinated a centralized digital pathology examination of the frozen tumor tissues collected for the study as well as FFPE sections when available, via a web-based report approach and dedicated expert panel, which consisted of seven experienced gastrointestinal pathologists. In brief, two frozen sections flanking the tumor tissues considered for DNA extraction were H&E stained and scanned by Leica digital slide scanner. High resolution images were randomly assigned to panel members, all blind to the original diagnosis. In addition to diagnosis and confirmation of tumor type, the percentage of viable cellular elements (tumor, inflammatory, and other non-tumor cells) and necrosis were recorded. A minimum of 50% viable tumor cells was required for eligibility to whole genome sequencing. In total, 1160 ESCC cases were enrolled into the study, 47.1% were excluded due to insufficient viable tumor cells (pathology level) or inadequate DNA (tumor or germline). Of the cases which proceeded to the final analysis (552), 95 (18.7%) cases were evaluated by two pathologists, among those, 78 (82%) were in full agreement. The main reason of disagreement in the remaining 17 evaluations was about the eligible cut off percentage of viable tumor cells. This was resolved by third independent evaluation to make a consensus.

### DNA extraction

Extraction of DNA from tumor and matched normal tissue was centrally conducted at IARC/WHO (collections from Brazil, China, Iran and West Africa), or at local centers (Japan and UK) following a similarly standardized DNA extraction procedure. Tumour DNA was extracted from fresh frozen material. Of the cases which proceeded to the final analysis (552), germline DNA was extracted from either buffy-coat (133), whole blood (382) or from adjacent normal tissue (37), using an Autopure (Qiagen) instrument as per the manufacturer’s instructions. Tumor DNA extraction was performed on the QIAsymphony (Qiagen) using the QIAsymphony DNA Mini Kit with the Tissue_HC DSP protocol according to the manufacturer’s instructions. When the amount of tumor tissue was limited (biopsy surface <10mm2), the Tissue_LC_DSP protocol was used instead. DNA quality was first assessed by spectrophotometry and quantifications were done using Quant-iT™ Picogreen ® dsDNA quantification reagent (Invitrogen).

### Whole genome sequencing

DNA from 613 cases was received at the Wellcome Sanger Institute for whole genome sequencing. Fluidigm SNP genotyping with a custom panel was performed to ensure that each pair of tumor and matched normal samples originated from the same individual. Whole genome sequencing (150bp paired end) was performed on the Illumina HiSeqX platform with target coverage of 40X for tumors and 20X for matched normal tissues. Whole genome sequencing data sequenced at an external center on the Illumina Novaseq platform was also available for 50 Japanese ESCC tumors with paired adjacent esophagus. The BAM files for externally sequenced case were converted to FASTQ and then analyzed using the same analysis pipeline as internally sequenced cases. All sequencing reads were aligned to the GRCh37 human reference genome using Burrows-Wheeler-MEM (v0.7.16a and v0.7.17). Cases were excluded if coverage was below 30X for tumor or 15X for normal tissue. Conpair (https://github.com/nygenome/Conpair) was used to detect contamination, cases were excluded if the result was greater than 3%^39^. A total of 552 cases passed all criteria and were included in subsequent analysis.

### Somatic Variant calling

Variant calling was performed using the standard Sanger analysis pipeline (https://github.com/cancerit). Copy number profiles were determined first using the algorithms ASCAT and BATTENBERG, where tumor purity allowed. SNV were called with cgpCaVEMan^40^, indels were called with cgpPINDEL^41^, and structural rearrangements were called using BRASS. CaVEMan and BRASS were run using the copy number profile and purity values determined from ASCAT where possible (complete pipeline, n=429). Where tumor purity was insufficient to determine an accurate copy number profile (partial pipeline, n=123), CaVEMan and BRASS were run using copy number defaults and an estimate of purity obtained from ASCAT/BATTENBERG. For SNV additional filters (ASRD >= 140 and CLPM ==0) were applied to remove potential false positive calls. To further exclude the possibility of caller specific artefacts being included in the analysis, a second variant caller, Strelka2, was run for SNVs and indels (Supplementary Methods)^42^. Only variants called by both the Sanger variant calling pipeline and Strelka2 were included in subsequent analysis.

### Germline variant calling

Germline variants were derived from the whole genome sequencing from the normal paired material for each individual using Strelka2 with appropriate quality control criteria^42^. Variant calls were then derived into genotypes for each individual and annotated using SNPeff. Genotypes for ALDH2 rs671, APOBEC rs12628403 polymorphisms were extracted for each individual, with individuals resolved into homozygote (ref) heterozygote (ref/alt) and homozygote (alt/alt). Population frequencies were consistent with that expected for these populations. For the BRCA1 and BRCA2 genes, SNPeff and clinvar annotations were used to identify variants that have a putative deleterious impact on the BRCA1 and BRCA2 gene products (Supplementary Table 18). CHORD (https://github.com/UMCUGenetics/CHORD) was run to predict homologous recombination deficiency in cases with germline BRCA1/2 variants.

### Generation of mutational matrices

The somatic mutations across all samples were transformed into five mutational matrices: SBS96, SBS288, DBS78, ID83, and SV32. Mutational matrices for single base substitutions (SBS), doublet base substitutions (DBS), and small insertions and deletions (ID) were generated using SigProfilerMatrixGenerator (https://github.com/AlexandrovLab/SigProfilerMatrixGenerator) with default options (v1.1.15)^43^. The SBS288 classification was introduced in the current manuscript as an alternative to the previously developed SBS384 classification^43^. Originally, SBS384 extended the traditional SBS96 scheme, which encompasses a single base substitution and its immediate 3’ and 5’ sequence context, by classifying each substitution as occurring: (i) on the transcribed strand (T); (ii) on the untranscribed strand (U); (iii) in a region of bidirectional transcription (B); or (iv) in intergenic/non-transcribed parts of the genome (N). In practice, very few mutations were classified in the B category. The SBS288 classification was created by removing the B mutational channels and splitting their mutations evenly into the appropriate transcribed (T) and untranscribed (U) mutational channels. The mutational matrix for structural rearrangements (SV32) was generated using SignatureToolsLib based on our previously developed classification in breast cancer^44, 45^.

### Mutational signature analysis

Mutational signatures were extracted using two algorithms, SigProfilerExtractor (https://github.com/AlexandrovLab/SigProfilerExtractor), based on non-negative matrix factorization, and HDP (https://github.com/nicolaroberts/hdp), based on the Bayesian hierarchical Dirichlet process. For SigProfilerExtractor, *de novo* mutational signatures were extracted from each mutational matrix using SigProfilerExtractor with default options (v1.0.14)^21^. Briefly, SigProfilerExtractor deciphers mutational signatures by first performing Poison resampling of the original matrix with additional renormalization (based on a generalized mixture model approach) of hypermutators to reduce their effect on the overall factorization^21^. Nonnegative matrix factorization (NMF) was performed using initialization with nonnegative singular value decomposition and by applying the multiplicative update algorithm using the Kullback–Leibler divergence as an objective function^21^. NMF was applied with factorizations between k=1 and k=25 signatures; each factorization was repeated 500 times and an optimum number of signatures was automatically detected by SigProfilerExtractor’s model selection method^21^. Where possible, SigProfilerExtractor matched each de novo extracted mutational signature to a set of previously identified COSMIC signatures^8^. HDP extraction of SBS96 signatures was performed without priors, using the country of origin to construct the hierarchy. SigProfilerExtractors’s decomposition module was subsequently used to match HDP *de novo* signatures to previously identified COSMIC signatures^8^.

### Attribution of activities of mutational signatures

The *de novo* (Sigprofiler) and COSMIC signature (Sigprofiler and HDP) activities were attributed for each sample using the MSA signature attribution tool https://gitlab.com/s.senkin/MSA46. For COSMIC attributions, only COSMIC reference signatures which were identified in the decomposition of *de novo* signatures where included in the panel for attribution. At its core, the tool utilizes the non-negative least squares (NNLS) approach minimizing the L2 distance between the input sample and the one reconstructed using available signatures. In order to limit false positive attributions, optimization procedure was applied by repeated removal of all signatures that do not increase the L2 similarity of a sample by > 0.02, a value suggested by simulations. Finally, a parametric bootstrap approach was applied in order to extract 95% confidence intervals for each attributed mutational signature activity.

### Driver mutations

Multiple approaches were combined to define driver mutations. Firstly, a dNdS approach was used to identify genes under positive selection in ESCC^22^. The analysis was preformed both for the whole genome (q<0.01), and with restricted hypothesis testing (RHT) for a panel of 369 known cancer genes^22^. Variants in any gene identified through the whole genome dNdS, any gene in the 369 cancer gene panel or in genes previously reported as significantly mutated in ESCC but not included either of the above criteria (*NOTCH3*, *FAT3*, *FAT4)* were assessed as potential drivers^13, 18^. Candidate driver mutations were annotated with the mode of action using the Cancer Gene Census (https://cancer.sanger.ac.uk/census) and the Cancer Genome Interpreter tool (https://www.cancergenomeinterpreter.org). Missense mutations were assessed using the MutationMapper tool (http://www.cbioportal.org/mutation_mapper). Variants were considered likely drivers if they met any of the following criteria:

- Truncating mutations in genes annotated as tumor suppressors
- Mutations annotated as likely or known oncogenic in MutationMapper
- Truncating variants in genes with selection (q <0.05) for truncating mutations assumed to be tumor suppressors and thus likely drivers
- Missense variants in all gene (excluding *C3orf70,* where hotspot mutations have been shown to be in DNA stems loops targeted by APOBEC)^47^ under positive selection and with dN/dS ratios for missense mutations above 5 (assuming 4 of every 5 missense mutations are drivers) labelled as likely drivers

### Clonality of APOBEC mutations

Clonality of APOBEC mutations was examined in cases with tumor purity >50% and with >0 mutations attributed to the APOBEC associated COSMIC reference signatures SBS2/SBS13. Median VAF was calculated for C>G and C>T mutations at TCN contexts and for all other SBS96 contexts. C>A mutations at TCN contexts were not considered due to the presence of SBS18.

### Clustered mutations

To identify clustered single base substitutions, a sample-dependent inter-mutational distance (IMD) cutoff was derived, which is highly unlikely to occur by chance given the mutational pattern and the number of mutations observed in each sample. Specifically, SigProfilerSimulator5 (v1.0.2) was used to simulate the random distribution of mutations in each sample that one would expect to observe by chance^48^. For each sample, SigProfilerSimulator was configured to maintain the +/− 2bp sequence context for each substitution, the total number of mutations within each chromosome, and the transcriptional strand bias ratios across all single base substitutions. All somatic mutations in each sample were simulated 100 times and an IMD cutoff was derived such as 90% of the mutations below this cutoff are clustered in a sample (i.e., not occurring purely by chance; q-value<0.01). Additionally, heterogeneity in mutation rates across the genome was also considered by correcting for mutation rich regions present in 10Mb-sized windows. Moreover, differences in clonality or copy-number were addressed using a threshold for the difference in variant allele frequencies between two subsequent substitutions (no more than 0.10) to ensure that a given subset of substitutions have occurred as a single event. Lastly, the identified clustered mutations were subclassified into specific categories of mutational events consisting of: (i) doublet substitutions; two adjacent mutations with consistent variant allele frequencies; (ii) extended multi-base substitutions, previously termed omikli events^49^, reflecting any two mutational events that are greater than 1bp and less than the sample-dependent IMD threshold with consistent variant allele frequencies; (iii) large mutational events, previously termed kataegis^50^, with three or more mutational events that are greater than 1bp and less than the sample-dependent IMD threshold with consistent variant allele frequencies.

### Principle component analysis

The VCF files containing the ESCC genotype data were converted into PLINK pgen format using PLINK (v2.00a; www.cog-genomics.org/plink/2.0/)^51^. Initially, ambiguous variants (8,352,843 out of 61,914,035) were removed. After performing basic quality control steps (missing genotype rates exceeding 10%, MAF < 1%, and Hardy-Weinberg equilibrium exact test p-value below 10^-6^) and considering only biallelic SNPs, 9,783,812 variants remained for further analyses. To detect and remove variants in linkage disequilibrium (LD), independent pairwise test (window size of 50 and r2 threshold of 0.2) was performed using PLINK and high LD regions (e.g. HLA region in chromosome 6) in reference genome (hg19) were also excluded from the analyses. The resulting 1,813,107 pruned variants were used to calculate the principal components (PC) in PLINK for the whole ESCC cohort (n=552, Extended Data Fig. 9a). The same procedure was applied for the cases from China and Japan (n=175) and 1,042,451 pruned variants were used to calculate the principal components for these countries (Extended Data Fig. 9b). For the associations between the germline variants and DNA mutational signatures, the first five principal components explaining about 55% of the variance in the dataset were included in the logistic regression models.

### Regressions

Epidemiological data for ESCC risk factors were collected using questionnaires from different retrospective studies. These data were harmonized in order to be used in common regression analysis across all regions. The harmonization was performed for indoor air pollution, family history of ESCC and temperature of food and drinks variables (Supplementary Table 20). Data on alcohol drinking and tobacco smoking were collected using standard questions in all five studies. Ever-smokers were defined as individuals who had smoked cigarettes, used hookah, or chewed tobacco at least weekly for 6 months or more. These individuals were further classified as current and former users, with current users defined as those who had practiced any of these habits within 1 year prior to the interview, and former users defined as those who had stopped their habit at least 1 year prior to the interview. Alcohol ever-drinkers were defined as individuals who had used any of commercially available or locally-made alcohol at least weekly for 6 months or more. Within the Iranian cohort, the general questionnaire covered opium use, which in Golestan includes teriak (crude opium), shireh (a refined opium extract) sukhteh (opium dross left in pipes after smoking opium), and heroin. Opium users were defined as subjects who had consumed opium at least once per week for a minimum of 6 months^52^. Signature attributions were dichotomized into presence and absence using confidence intervals, with presence defined as both lower and upper limits being positive, and absence as the lower limit being zero. An exception was made for COSMIC signature SBS3 as this signature exhibited wide confidence intervals mostly consistent with zero. According to simulations, SBS3 attribution remained highly specific (specificity >95%) without application of confidence intervals, so any positive attribution was treated as presence of SBS3, and absence otherwise. The binary attributions served as dependent variables in logistic regressions, and relevant risk factors were used as factorised independent variables. In order to adjust for confounding factors, sex, age of diagnosis, region, as well as alcohol and tobacco statuses were added as covariates in regressions. The region variable was categorized as high incidence (>20/100,000; China, Iran, Kenya, Malawi, Tanzania) or low incidence (<6/100,000; Brazil, Japan, UK). Regressions with signatures SBS16 and ID11 with respect to alcohol status and region were performed using the penalized logistic regression (Firth method) since these signatures were exclusively attributed in ever-drinkers, with the exception of one case with ID11. For a similar reason of data separation, SBS288I regression with tobacco smoking was performed using Firth method. Mixed models with random intercept, with and without random effects for exposure were attempted where possible, yet did not provide any benefit, yielding very similar results (Supplementary Table 21).

Regressions with respect to the germline variants data were performed in a similar fashion using logistic regressions. Confounding factors used in these regressions were limited to sex, age of diagnosis and genetic ancestry determined using principal component analysis (PCA). Regressions with rs671 germline variant genotype were limited to China and Japan since this variant was only observed in these populations. Genetic ancestry principal components were removed as covariates as neither of alcohol related signatures SBS16 and ID11 were present in the Chinese cohort, perfectly separated from the Japanese cohort by the first principal component (see Extended Data Fig. 9). Cases with the rs11571833 (BRCA2) ESCC risk allele were excluded for regressions with BRCA1/2 germline variants as this allele has been classified as likely benign in clinvar.

## Data availability

Whole genome sequencing data and patient metadata are deposited in the European Genome 715 Phenome Archive (EGA) associated with study EGAS00001002725. BAM files for all cases included in the final analysis were deposited in dataset EGAD00001006868, and patient metadata in dataset EGAD00001006732. All other data is provided in the accompanying Supplementary Tables.

## Code availability

All algorithms used for data analysis are publicly available with repositories noted within the respective method sections and in the accompanying reporting summary. Code used for regression analysis is available at https://gitlab.com/Mutographs/Mutographs_ESCC. Any other code is available on request.

## Acknowledgements

The authors would like to thank Laura O’Neill, James Hewinson, Kirsty Roberts, Nathalie Smerdon, James Mack, Emma Gray and the staff of DNA Pipelines at the Wellcome Sanger Institute for their contribution. We are grateful for the support provided by the IARC ESCCAPE team (Margaret Odour, Nicholas Kigen, Fatma Some (Eldoret), Godfrey Mushi (Moshi), Mary Suwedi, Thandiwe Soloman, Rose Malamba Blantyre and Daniel Middleton), Sandrine Magat, Philippe Boutarin, Daniel Middleton and Atieh Hajimohammadsadegh, as well as IARC General Services, including the Laboratory Services and Biobank team led by Zisis Kozlakidis, the Section of Support to Research overseen by Tamas Landesz and the Evidence Synthesis and Classification Section led by Ian Cree, under IARC regular budget funding. The authors would like to thank Carol Giffen, Phil Taylor, and Amy Hutchinson for help with data/sample preparation and processing. The authors would like to thank Peter Campbell, Inigo Martinocorena, Tim Butler, Luiza Moore, Daniel Leongamornlet, Philip Robinson, Tim Coreens, Adi Steif, Burcak Otlu, Christophe Lallemand, Helene Renard, Theodore Cholin, Priscila Chopard, Maxime Vallee, Maja Milojevic, Maggie Blanks and Mimi McCord for useful discussions. The authors would also like to thank all the patients involved in this study.

## Funding

This work was supported by a Cancer Grand Challenges Mutographs team award funded by Cancer Research UK [C98/A24032] and Wellcome grant reference 206194. The work was also partly funded by R21CA191965 (Eldoret), Wereld Kanker Onderzoek Fonds (WKOF) / World Cancer Research Fund (2018/1795) and IARC Section of Environment and Radiation. The work of R.C.C.P. reported in this paper was undertaken during the tenure of an IARC Postdoctoral Fellowship at the International Agency for Research on Cancer. The laboratory of R.C.F. is funded by a Core Programme Grant from the Medical Research Council (RG84369), this research was supported by the NIHR Cambridge Biomedical Research Centre (BRC-1215-20014) and OCCAMS2 was funded by a Programme Grant from Cancer Research UK (RG81771/84119).

The work was also partly funded by the Practical Research Project for Innovative Cancer Control from the Japan Agency for Medical Research and Development (AMED) (JP20ck0106547h0001 to T.S.). A.M.G. and N.H were supported by the Intramural Research Program of the Division of Cancer Epidemiology and Genetics, National Cancer Institute, National Institutes of Health. L.B.A is an Abeloff V Scholar and is supported by an Alfred P. Sloan Research Fellowship. Research at UC San Diego was also supported by a Packard Fellowship for Science and Engineering to L.B.A.

## Author contributions

Analysis of data was performed by S.M., S.S., S.M.A.I., J.W., D.N., R.C.C.P., S.F., E.N.B., J.A., Y.H., A.K., V.G., C.L., E.T., I.A., P.E.B., D.J., J.W.T. Pathology review was carried out by B.A-A., S.S., J-Y.S., H.S., F.A-A., M.S., A.N., M.E., P.R., L.C. DNA extraction was carried out by C.C. Patient and sample recruitment was led by R.C.F., L.F.P., C.D., B.T.M., D.M., A.M.G., N.H., R.M., A.F., V.M. Patient and sample recruitment and sequencing for Japanese cases was led by T.S. Scientific Project Management was carried out by L.H., E.C., G.S. and S.P. S.M. and S.S. jointly contributed and were responsible for overall scientific coordination. The manuscript was written by S.M. and M.R.S. with contributions from all other authors.

## Competing interests

The authors declare no competing interests.

## Disclaimer

Where authors are identified as personnel of the International Agency for Research on Cancer or the World Health Organization, the authors alone are responsible for the views expressed in this article and they do not necessarily represent the decisions, policy, or views of the International Agency for Research on Cancer or the World Health Organization.

## Data Availability

https://ega-archive.org/studies/EGAS00001002725

---

**Extended Data Fig.1:**
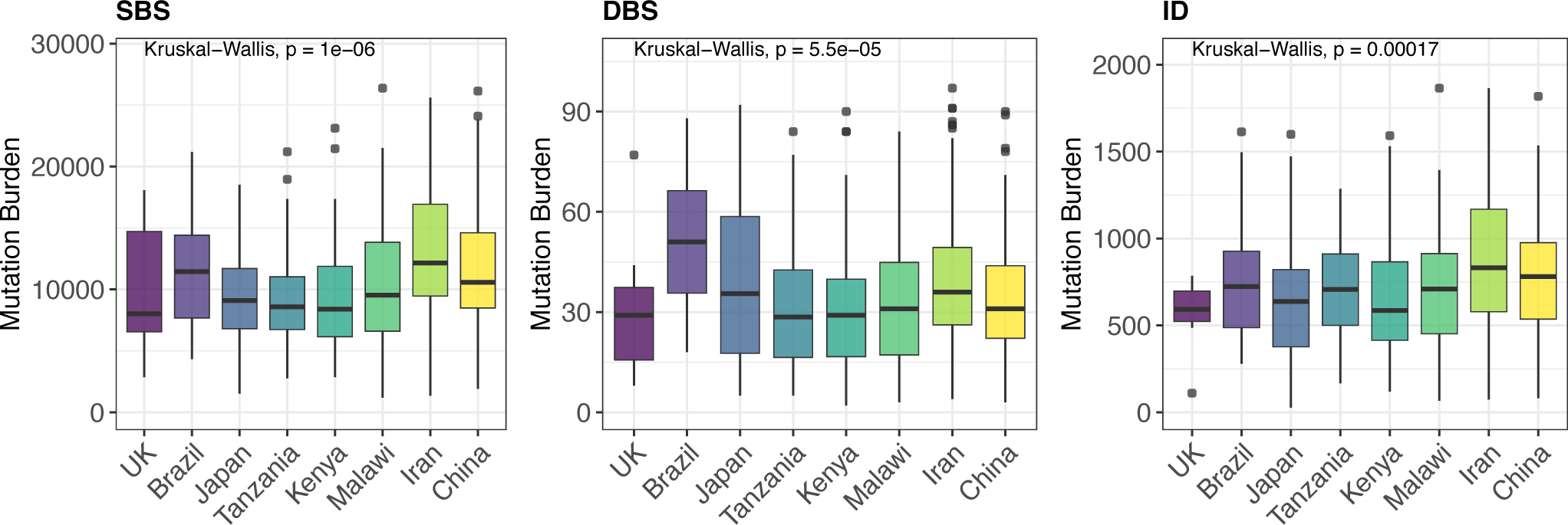
Mutation burdens in ESCC. Mutation burdens for SBS, DBS and ID variants show modest differences between countries. Hypermutators in each group were defined using the interquartile range method, with cases with mutation burdens more than 1.5 IQR above Q3 removed from the analysis. Box and whiskers plots are in the style of Tukey. The line within the box is plotted at the median while the upper and lower ends are indicated 25th and 75th percentiles. Whiskers show 1.5*IQR (interquartile range) and values outside it are shown as individual data points. Countries are ordered by approximate incidence rate in ascending order.

**Extended Data Fig.2:**
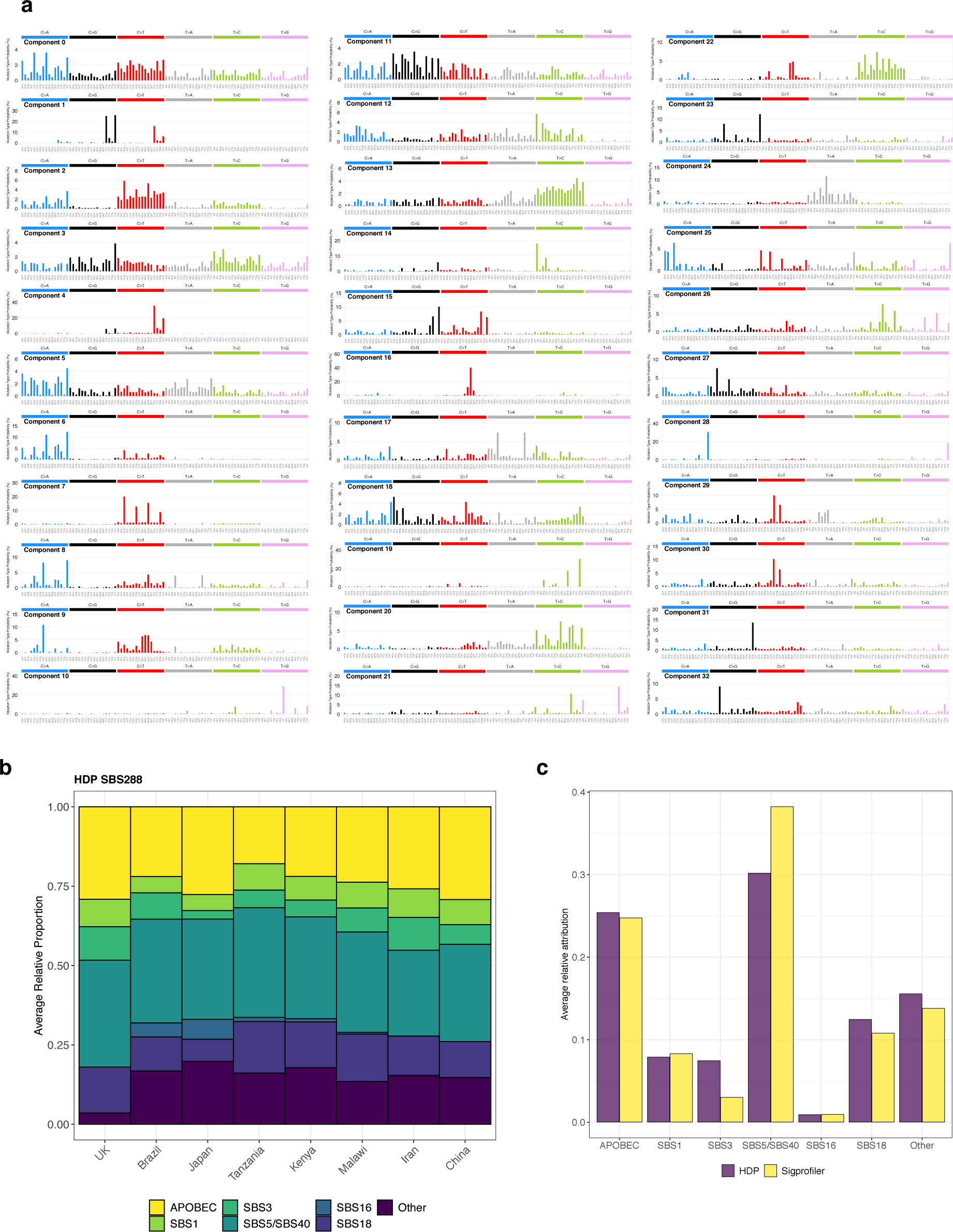
HDP mutational signature analysis of ESCC. (a) 33 components extracted by HDP. (b) Average relative attributions of SBS COSMIC signatures and SV *de novo* signatures are broadly similar between all countries. Signatures accounting for less than 5% on average (with the exception of SBS3 and SBS16) are grouped together into the ‘Other’ category. (c) Comparison of average relative attribution of signatures between HDP and Sigprofiler.

**Extended Data Fig.3:**
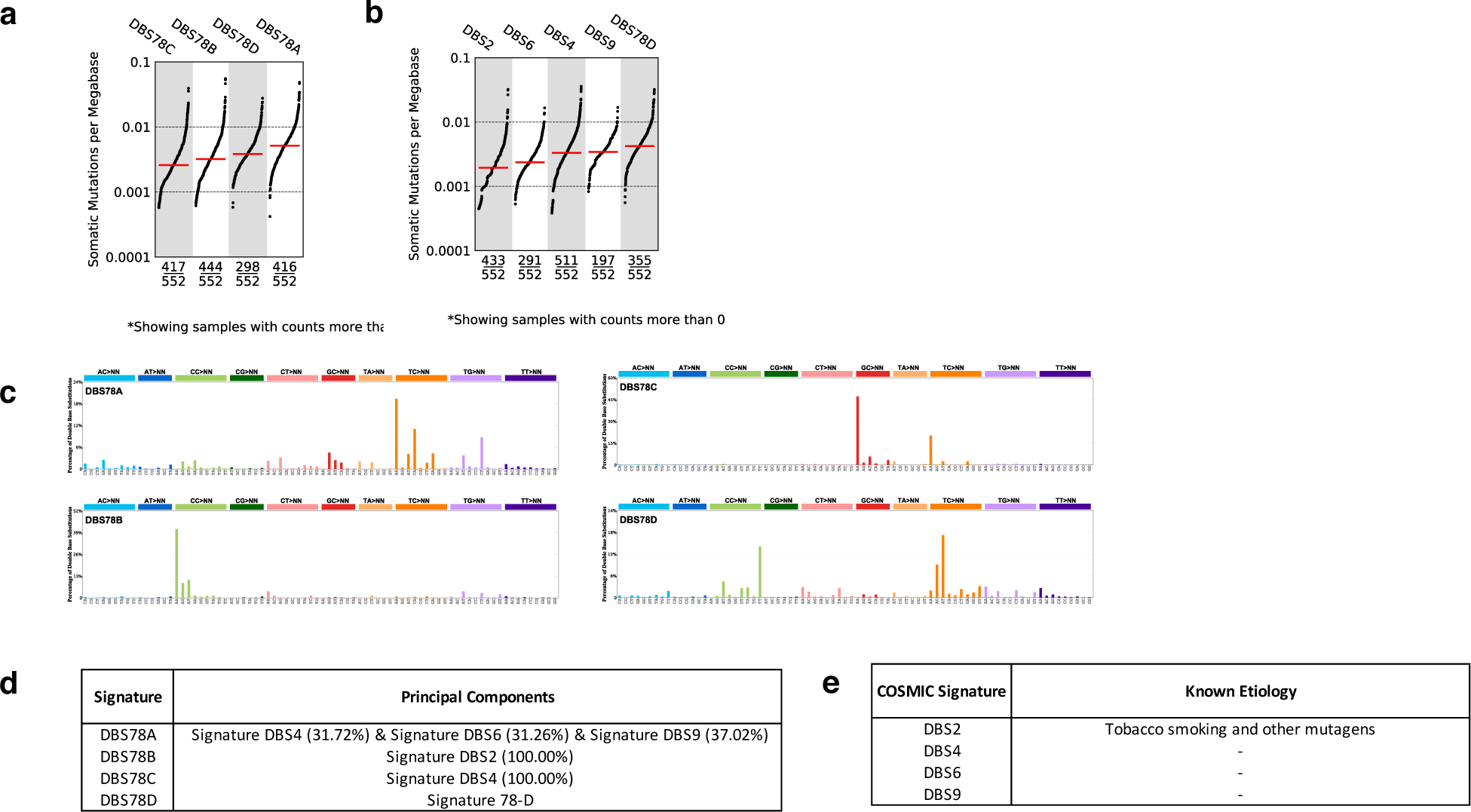
DBS78 mutational signature analysis of ESCC. (a) TMB plot showing the frequency and mutations/mb for each of the extracted DBS78 *de novo* signatures. (b) TMB showing the frequency and mutations/mb for each DBS78 COSMIC reference signature identified in the ESCC cohort. (c) Four *de novo* DBS78 signatures extracted from 552 ESCC cases. (d) Mapping of de novo signatures extracted from ESCC decomposed into COSMIC reference signatures. (e) COSMIC reference signatures identified in the ESCC cohort with known etiologies.

**Extended Data Fig.4:**
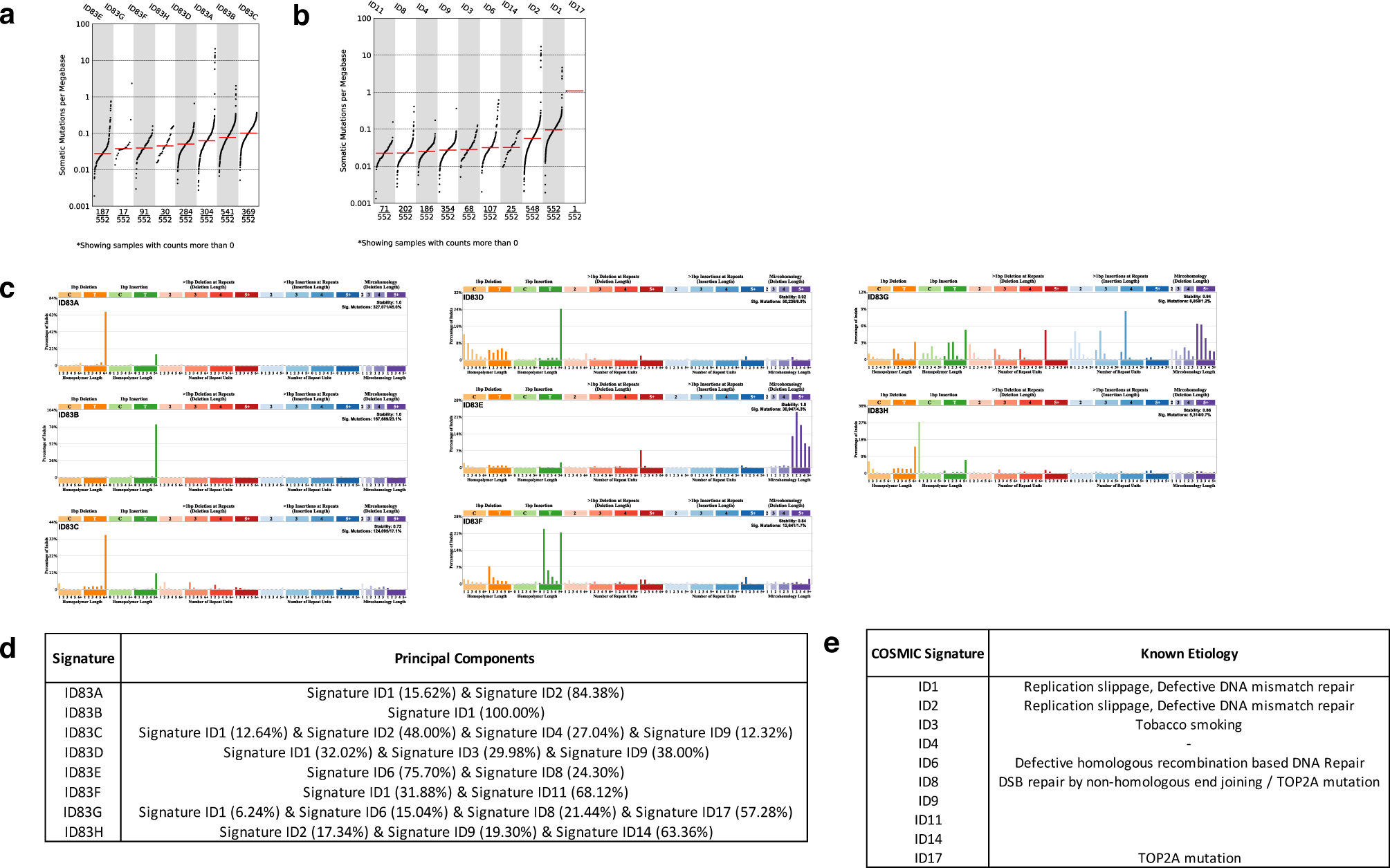
ID83 mutational signature analysis of ESCC. (a) TMB plot showing the frequency and mutations/mb for each of the extracted ID83 *de novo* signatures. (b) TMB showing the frequency and mutations/mb for each ID83 COSMIC reference signature identified in the ESCC cohort. (c) Eight *de novo* ID83 signatures extracted from 552 ESCC cases. (d) Mapping of de novo signatures extracted from ESCC decomposed into COSMIC reference signatures. For clarity, components amounting to less than 10% of the signature have been omitted. (e) COSMIC reference signatures identified in the ESCC cohort with known etiologies.

**Extended Data Fig.5:**
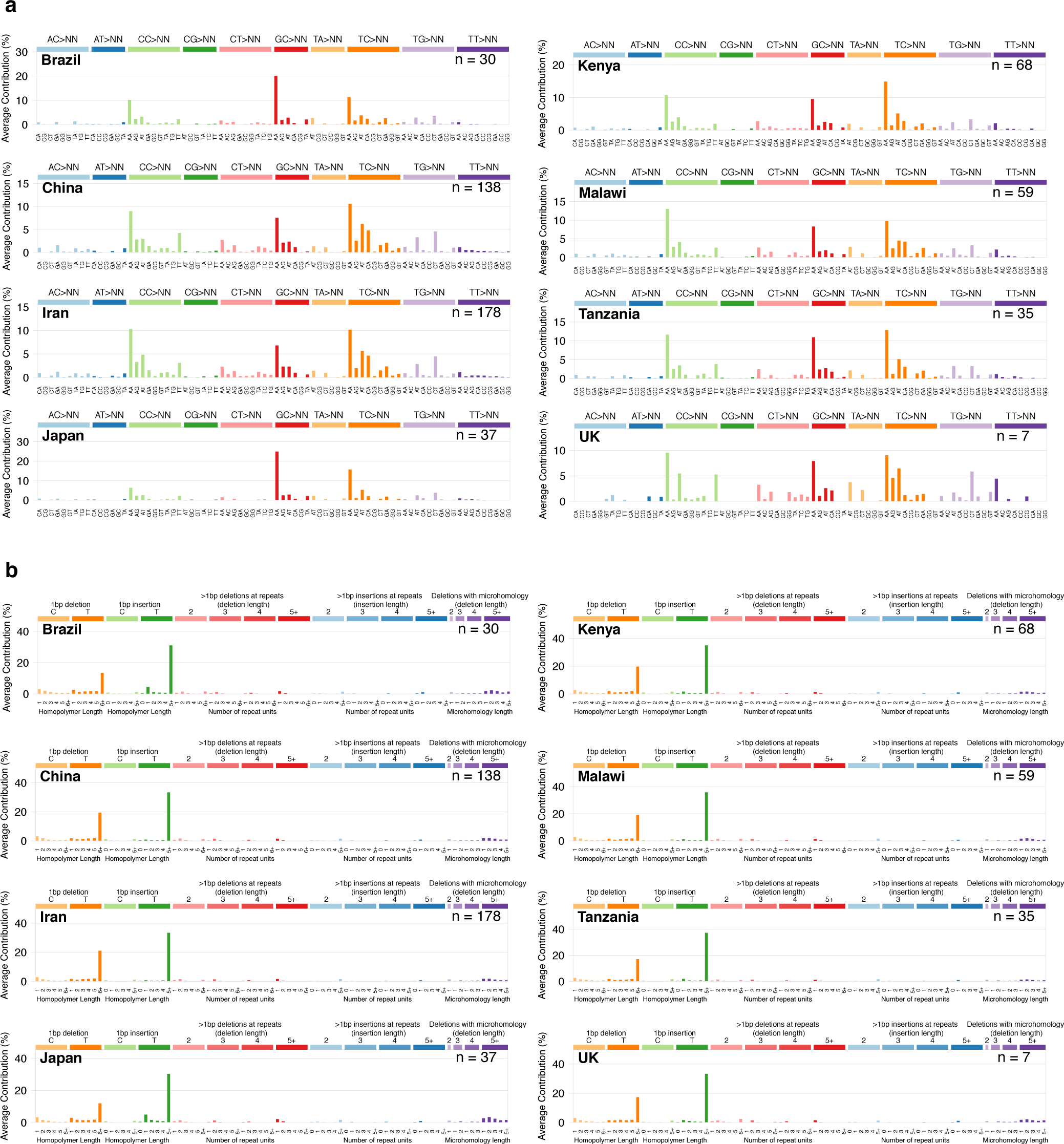
Average mutational spectra of ESCC from eight countries. (a) Average DBS78 spectra in ESCC from eight countries (b) Average ID83 spectra in ESCC from eight countries.

**Extended Data Fig.6:**
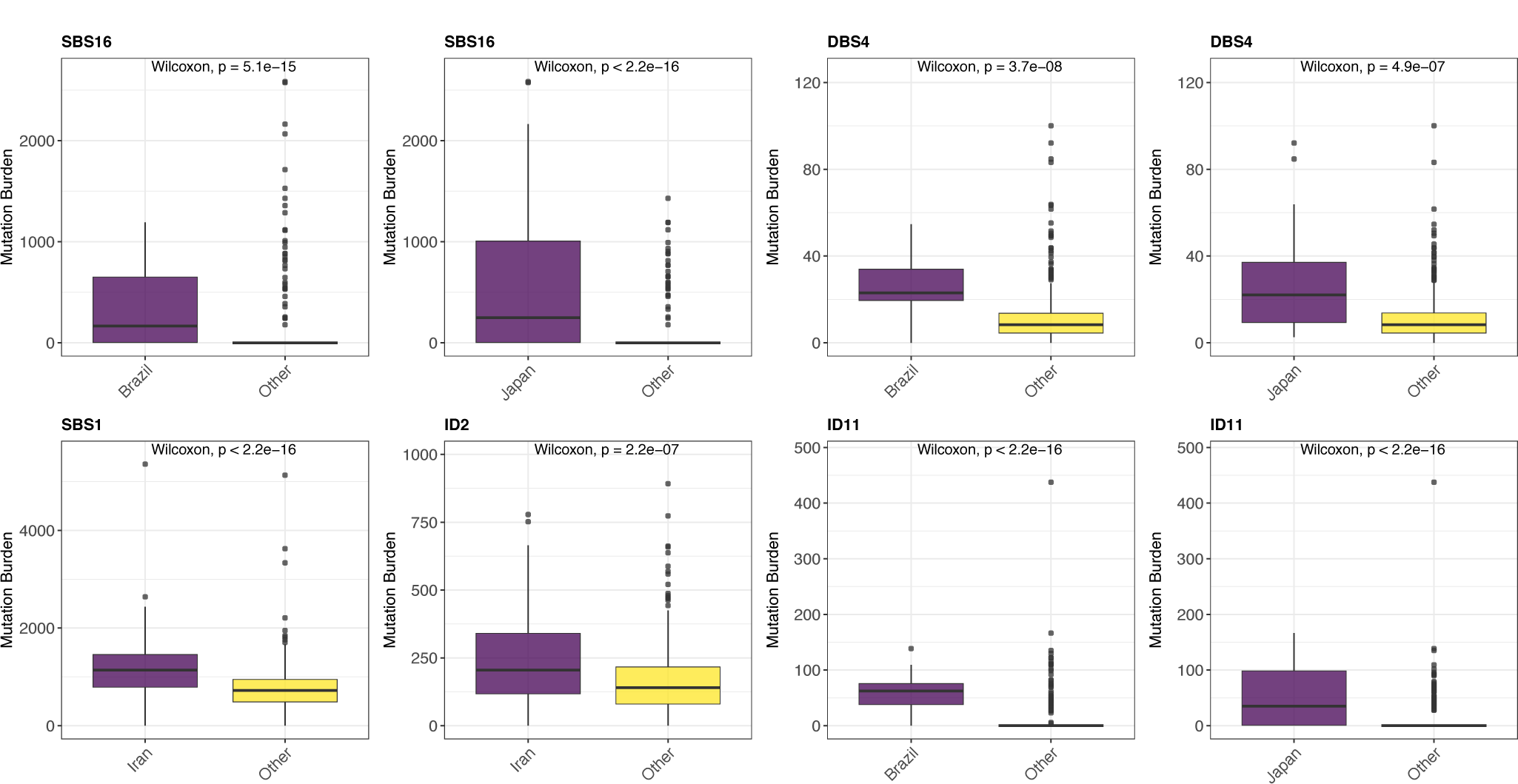
Differences in mutation signature mutation burden in ESCC. Countries showing significant differences in attributed mutation burden of mutational signatures when grouped against all other countries. The Wilcoxon signed-rank test (two-sided) was used to test for significant differences, for signatures where a significant global difference was found using the Kruskal-Wallis test with multiple hypothesis corrections (Fig2). For clarity, cases with >1000 mutations attributed to ID2 are not displayed. Box and whiskers plots are in the style of Tukey. The line within the box is plotted at the median while the upper and lower ends are indicated 25th and 75th percentiles. Whiskers show 1.5*IQR (interquartile range) and values outside it are shown as individual data points.

**Extended Data Fig.7:**
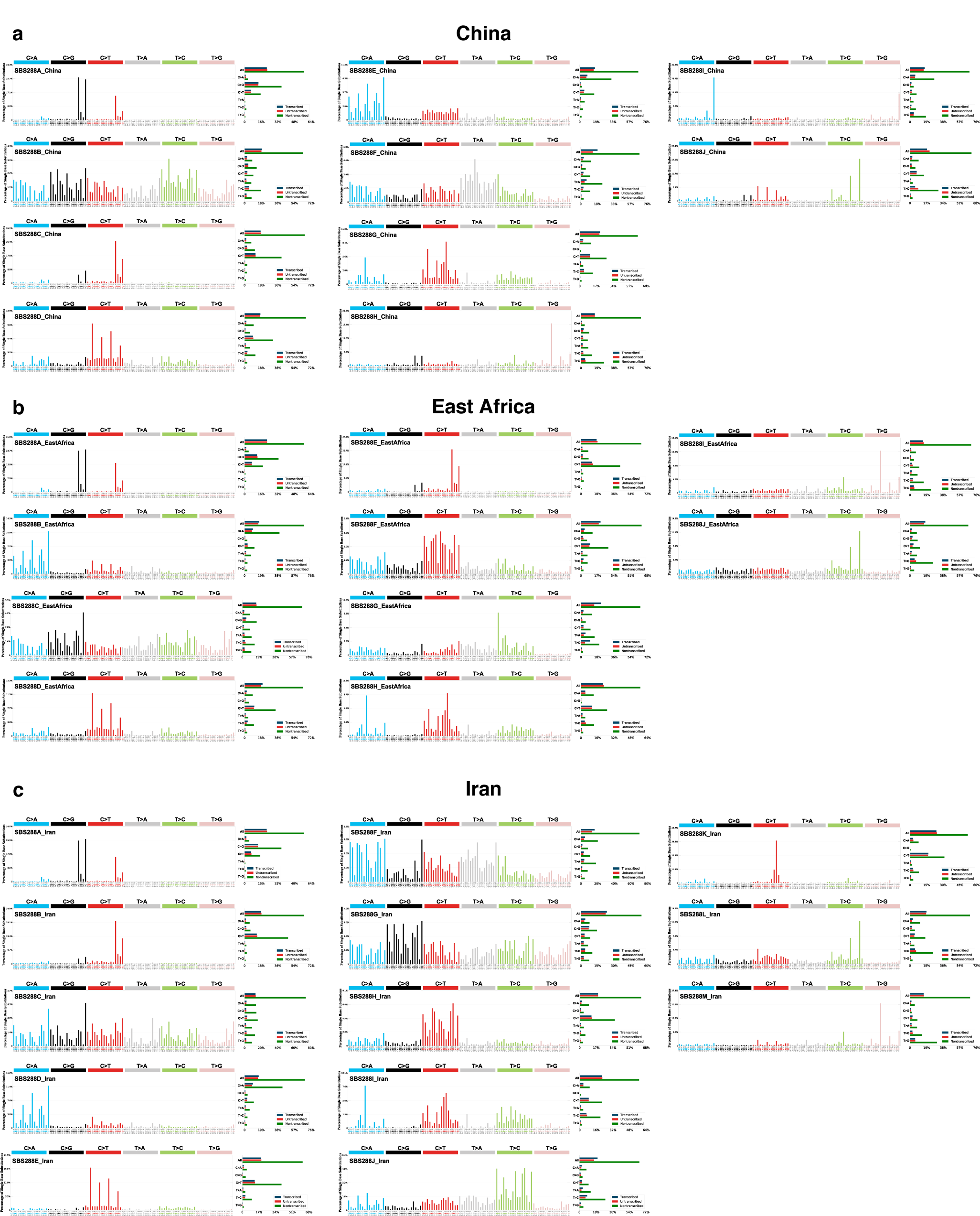
Regional SBS288 signature extractions. (a) Ten *de novo* signatures extracted from 138 ESCC cases from Shanxi province, China. (b) Ten *de novo* signatures extracted from 162 ESCC cases from Kenya, Tanzania and Malawi. (c) Thirteen *de novo* signatures extracted from 178 ESCC cases from Golestan province, Iran.

**Extended Data Fig.8:**
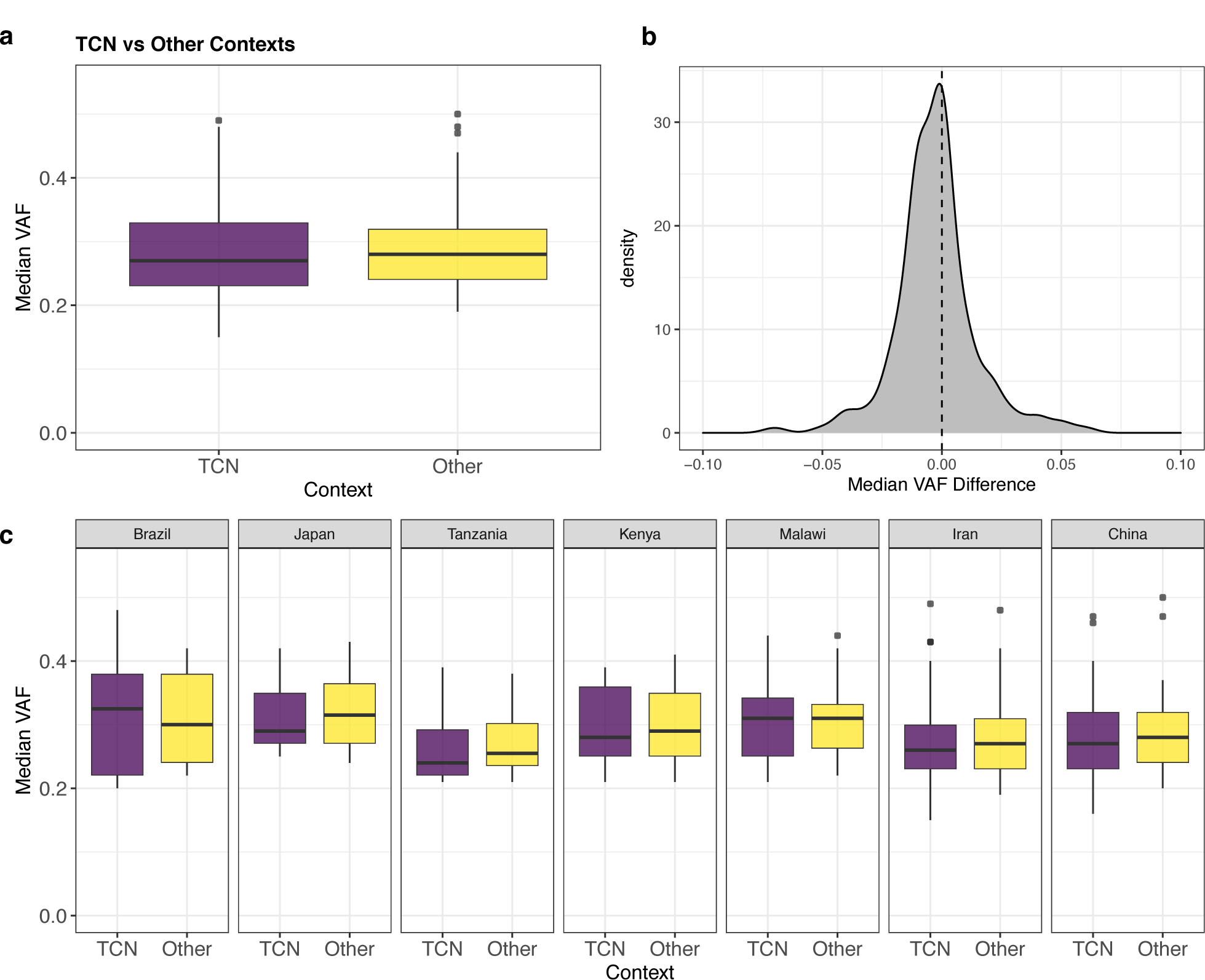
Comparison of VAF at TCN and other contexts. (a) C>G and C>T variants at TCN contexts have a similar median VAF to those at other contexts. Cases with tumor purity <50% and those with no attribution of APOBEC mutational signatures (COSMIC signatures SBS2 and SBS13) were excluded from the analysis. Box and whiskers plots are in the style of Tukey. The line within the box is plotted at the median while the upper and lower ends are indicated 25th and 75th percentiles. Whiskers show 1.5*IQR (interquartile range) and values outside it are shown as individual data points. (b) There is no shift in the median VAF (Median VAF(TCN) – Median VAF (other contexts)) of C>G and C>T variants at TCN contexts compared to variants at other contexts. Cases with tumor purity <50% and those with no attribution of APOBEC mutational signatures (COSMIC signatures SBS2 and SBS13) were excluded from the analysis. (c) No significant difference was found in the median VAF of C>G and C>T variants at TCN contexts compared to all other contexts in any individual country. UK cases were not included as only two cases were of sufficient purity. Countries are ordered by approximate incidence rate in ascending order.

**Extended Data Fig.9:**
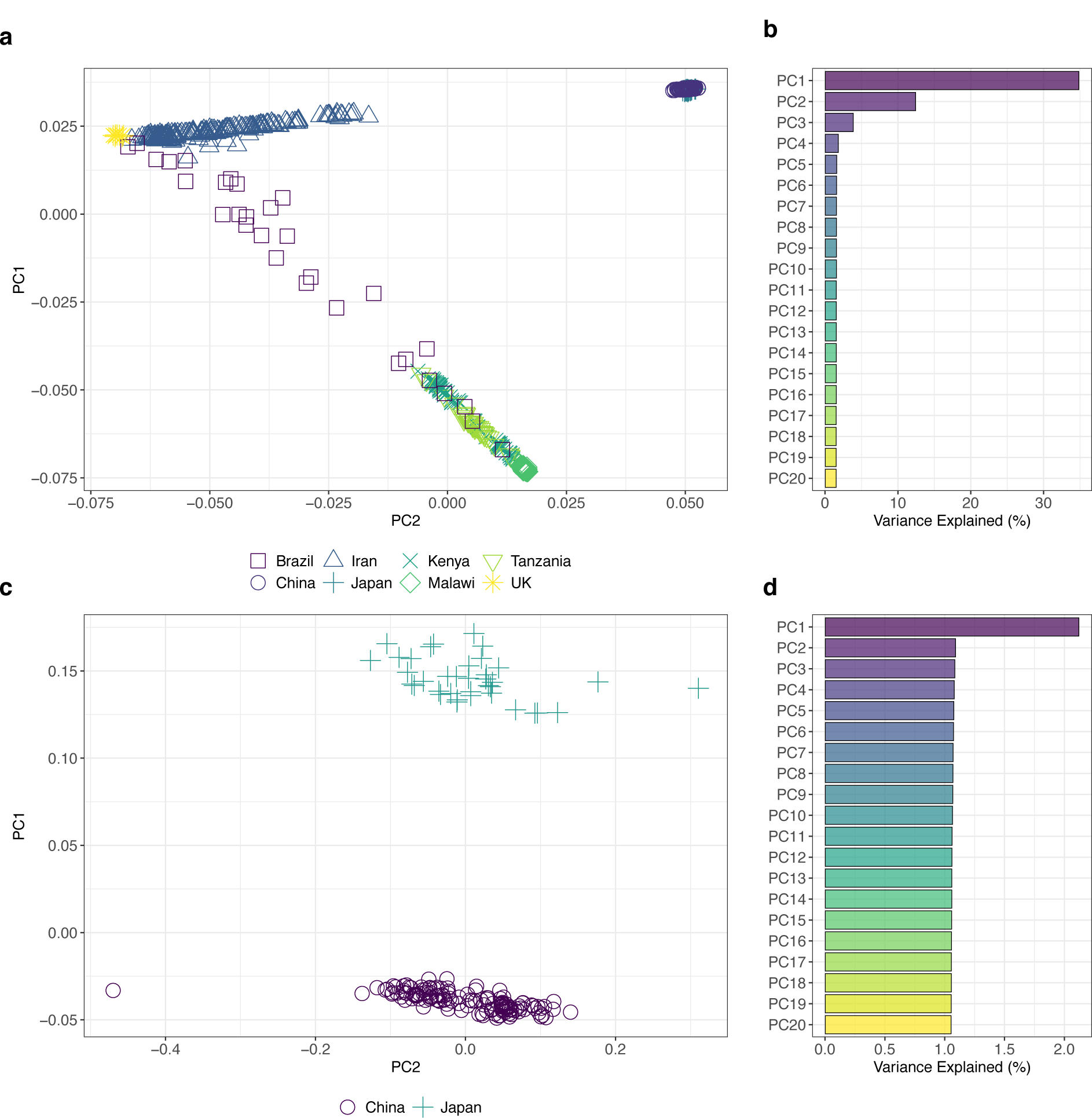
Genetic population structure based on principle component analysis. (a) Scatter plots of principal components PC1 and PC2 based on genotype data showing the genetic structure of the ESCC cohort across different countries of origin. (b) Bar plot showing the percentage of explained variance by each principal component (eigenvalue in percent) across all countries of origin (c) Scatter plots of principal components PC1 and PC2 based on genotype data showing the genetic structure of the ESCC cases from China and Japan. (b) Bar plot showing the percentage of explained variance by each principal component in ESCC cases from China and Japan (eigenvalue in percent).

## Notes

### Competing Interest Statement

The authors have declared no competing interest.

### Author Declarations

Ethical approvals were first obtained from each Local Research Ethics Committee and Federal Ethics Committee when applicable, as well as from the IARC Ethics Committee (IRB number: IEC Project 17-10)

