## Supplementary Methods for "Mutational signatures in esophageal squamous cell carcinoma from eight countries of varying incidence"

#### Comparison of Mutational Patterns from Different Variant Calling Approaches

In this study we used the combination of CaVEMan + Strelka2 and Pindel + Strelka2 for the final set of single base substitutions and indels. In order to examine whether the choice of second caller had a significant impact on the mutational profiles, 10 evaluated samples were compared with different variant call approaches (Supplementary Methods Fig.1), including GATK4 Mutect2[1], Strelka2[2], Varscan2[3] and MuSE[4]. Note that MuSE only identifies single base substitutions while the other three tools also provide small insertions and deletions. For Mutect2, paired reads were allowed to independently support different haplotypes during initial variant calling and the expected frequency of alleles not found in the germline resource was 0.00003125 (considering 16,000 as the number of protein coding genes known in the human genome). Contamination table and read orientation models were built from the paired samples and were subsequently used for filtering. For VarScan2, the initial variant calling expected a tumor purity of 0.8 and the subsequent filtering required a minimum coverage of 10 reads and at least 3 alternative reads in tumor with a minimum alternative allele frequency of 0.20. For Strelka2 and MuSE, the default setting for whole genome sequencing were used to produce a list of raw and filtered variants. Cosine similarities between the SBS96 contexts of single base substitutions were calculated by comparing the reference to mutations from (i) Mutect2, (ii) Strelka2, (iii) Varscan2, (iv) MuSE, (v) CaVEMan + Mutect2, (vi) CaVEMan + Varscan2, (vii) CaVEMan + MuSE. Similarly, cosine similarities between the ID83 contexts of small insertions and deletions were calculated by comparing the reference to mutations from (i) Mutect2, (ii) Strelka2, (iii) Varscan2, (iv) CaVEMan + Mutect2, (v) CaVEMan + Varscan2. The results showed that regardless of the second caller used, the cosine similarities of the mutational spectra generated were very similar (Supplementary Methods Fig.1).

**Supplementary Methods Figure Legend**

**Supplementary Methods Fig 1. Comparison of Mutational Patterns from Different Variant Calling Approaches.** An example comparing the patterns of somatic mutations for a single ESCC case is shown for both single base substitutions (a) and small insertions and deletions (b). The mutational profile of single base substitutions are highly stable across different variant callers in this sample (a) as well as across all 10 examined samples (c). Specifically, when compared to the reference, Mutect2, MuSE and all 2-caller approaches produced cosine similarity > 0.99 while Strelka2 and Varscan2 each showed patterns with median cosine similarity > 0.95 (c). The mutational profile of small insertions and deletions were less stable in this case and across all other examined cases. When compared to the reference, Mutect2, Strelka2 and Varscan2 generated mutational patterns with median cosine similarities between 0.92 and 0.97. On the contrary, MuSE and all of the 2-callers yielded highly similar spectrums (cosine similarity close to 1).

### Supplementary Methods Fig 1. Comparison of Mutational Patterns from Different Variant Calling Approaches

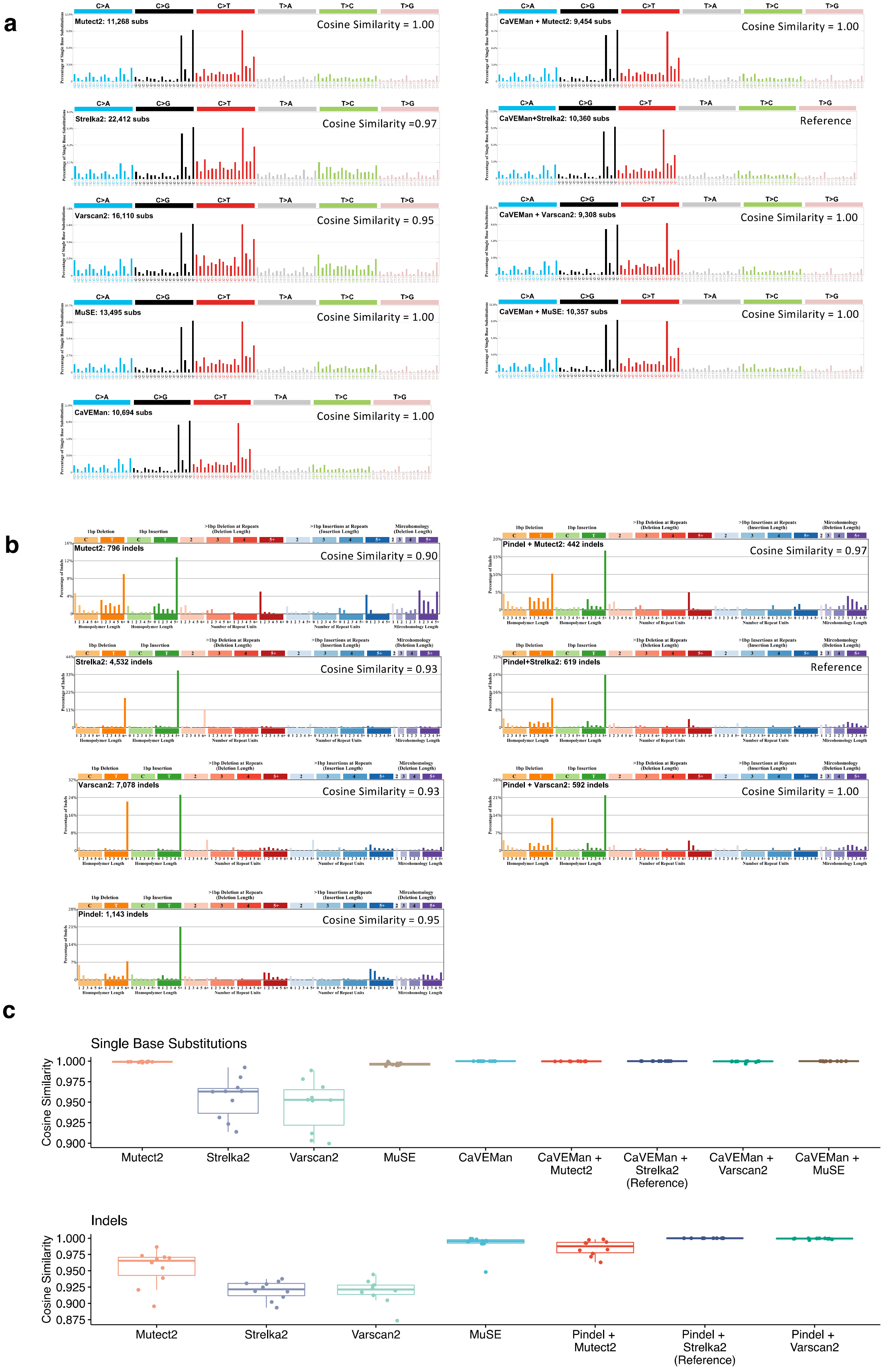
