## Supplementary Results for "Mutational signatures in esophageal squamous cell carcinoma from eight countries of varying incidence"

#### Copy Number Profiles

Whilst this dataset was not ideal to investigate copy number changes as the cohort is comprised of cases with variable purity, which impacts the reliability of copy number profile calling, the copy number profiles were investigated in a subset of cases with tumour purity >50%. As expected, overall the copy number profiles in ESCC were complex, with no significant differences in the number of CNV segments detected (Supplementary Results Fig. 1, Supplementary Table 1).

**Supplementary Results Fig.1: Copy number profiles in ESCC**

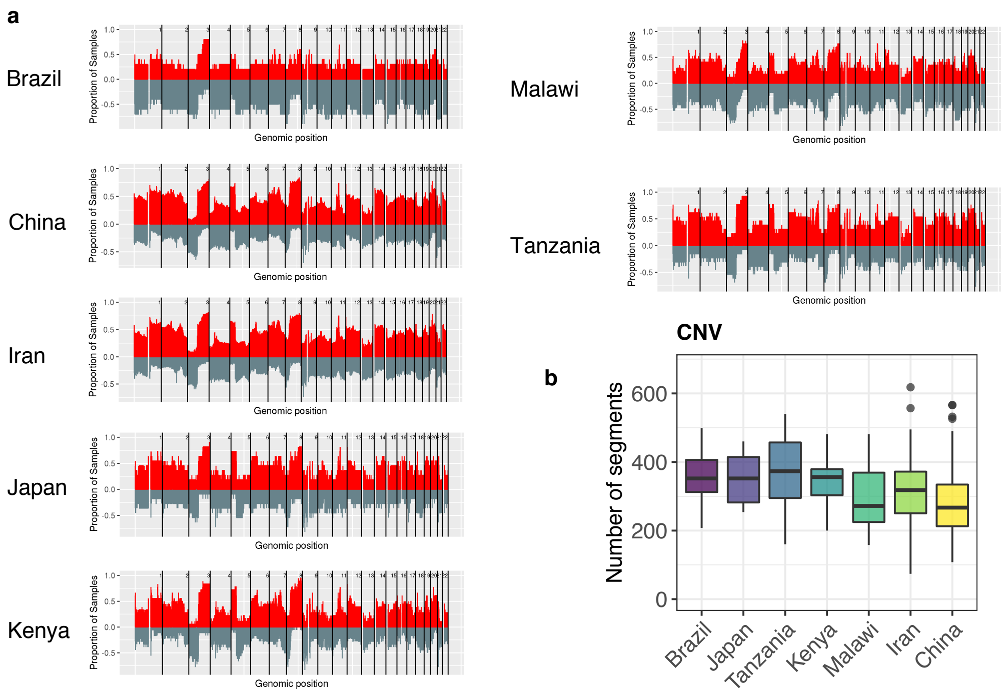


**Supplementary Results Fig.1: Copy number profiles in ESCC.** (a) Complex patterns of gains and loss in 185 ESCC tumor genomes. Cases with tumor purity <50% were excluded from this analysis, and UK cases were not included as only two cases were of sufficient purity. (b) Number of CNV segments per case in 185 ESCC tumor genomes as determined by the ASCAT algorithm, showing no significant difference between countries. Box and whiskers plots are in the style of Tukey. The line within the box is plotted at the median while the upper and lower ends are indicated 25th and 75th percentiles. Whiskers show 1.5*IQR (interquartile range) and values outside it are shown as individual data points. Countries are ordered by approximate incidence rate in ascending order.

### Novel *de novo* SBS and DBS signatures

One SBS288 *de novo* signature (SBS288P) could not be decomposed into COSMIC reference signatures, and was found in 30/552 (5%) cases. This signature was present in the HDP results (Extended Data Fig. 3a) and has also been identified in stomach cancers from a recent reanalysis of the pan cancer cohort used to derive the COSMIC reference signatures, but the etiology remains unknown[1]. SBS288P was most frequent in China (15/138, 11%), followed by Malawi (3/59, 5%), Kenya (3/68, 4%), Iran (8/178, 4%) and Brazil (1/30, 1%), but was not observed in Japan, Tanzania or the UK, most likely due to the small size of these cohorts.

The novel DBS *de novo* signature DBS78D was present in 64% of cases with an average contribution of 26% to the mutation burden. This signature could not be decomposed, but contained CC>NN peaks consistent with COSMIC reference DBS11, which is thought to be associated with APOBEC mutagenesis, in addition to TC>NN peaks which whilst present in DBS11, were more prominent in the novel extracted signature[2].

### Structural rearrangement signatures

The number of rearrangements ranged from 1 – 1110 (median 218) per case, with no differences between countries observed (Supplementary Results Fig. 2a). Structural rearrangement signatures were extracted using the 32 subclasses previously described[3]. This resulted in ten *de novo* signatures (Supplementary Results Fig. 2b-d, Supplementary Table 5,8), which could not be further decomposed as reference rearrangement signatures do not currently exist. The dominant signature was RS32A, which was defined by non-clustered translocations. This signature was present in 98% of cases and on average accounted for 33% of the rearrangement burden. The remaining signatures accounted for between 4-11% of the rearrangement burden on average. RS32A, RS32B and RS32I were similar to breast cancer rearrangement signatures RS2, RS4 and RS1 respectively (Supplementary Table 7), whereas the other signatures did not closely resemble those previously reported[3]. No significant differences in the attributions of any of the rearrangement signatures was observed in any individual country. (Supplementary Results Fig. 2c, Supplementary Tables 8, 10).

Rearrangement signatures were included in the regression analysis; however, no signatures were found between and ESCC risk factors and any of the *de novo* signatures. Despite previous studies finding associations between cases with BRCA1/2 variants and structural rearrangement signatures in breast cancer, no significant associations were found in ESCC[3].

**Supplementary Results Fig.2: SV32 mutational signature analysis of ESCC.** (a) No significant difference was observed in the number of rearrangements between countries. Hypermutators were defined using the interquartile range method, with cases with mutation burdens more than 1.5 IQR above Q3 removed from the analysis. Box and whiskers plots are in the style of Tukey. The line within the box is plotted at the median while the upper and lower ends are indicated 25th and 75th percentiles. Whiskers show 1.5*IQR (interquartile range) and values outside it are shown as individual data points. Countries are ordered by approximate incidence rate in ascending order. (b) TMB plot showing the frequency and mutations/mb for each of the extracted SV32 *de novo* signatures. (c) Average relative attributions of SV *de novo* signatures are broadly similar between all countries. Countries are ordered by approximate incidence rate in ascending order. (d) Ten *de novo* signatures extracted from 552 ESCC cases.

### Supplementary Results Fig.2: SV mutational signature analysis of ESCC

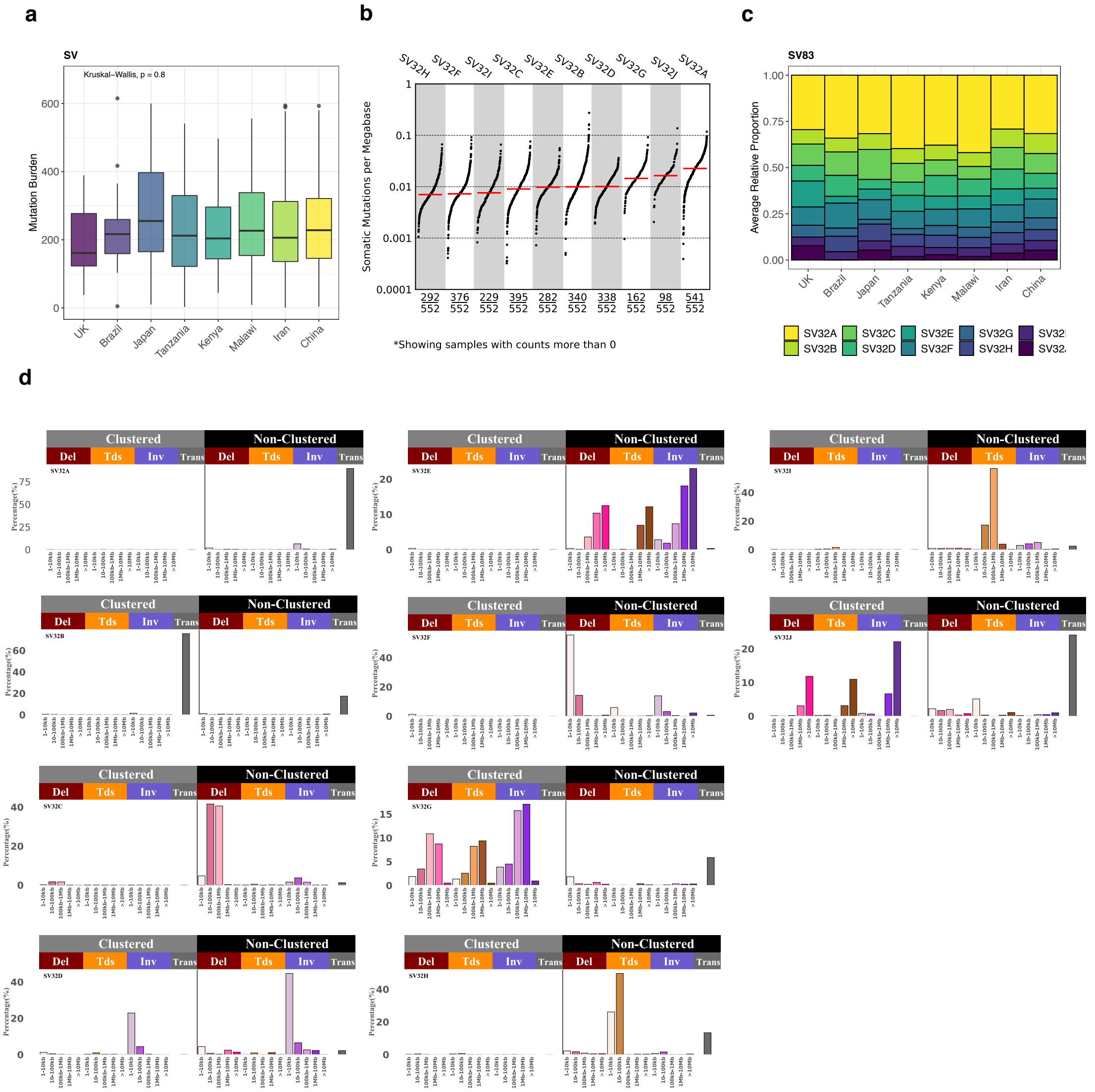


### Clustered mutations

The number and type of clustered mutations were investigated in the subset of cases with tumor purity >50% (Methods). This included doublet substitutions, the recently defined omikli events[4] and kataegis events, which collectively represented an average 3.24% (0.82%, 1.96% and 0.46% respectively) of the total SBS mutation burden. There was no significant difference between countries in either the total number or proportion of any clustered mutation type (Supplementary Results Fig.3).

**Supplementary Results Fig.3: Clustered mutations in ESCC**

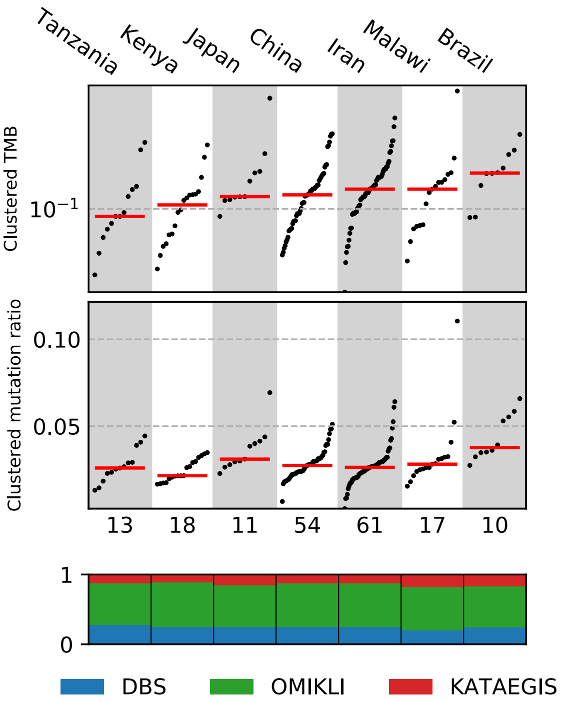


**Supplementary Results Fig.3: Clustered mutations in ESCC.** The distribution of clustered mutations in ESCC compared across each country ordered by median tumor mutational burden, where each dot represents a single tumor. The clustered mutation ratio is calculated as the fraction of clustered mutations compared to the total number of mutations in a given sample. Each clustered event is subclassified and summarized as the proportion of mutations per country associated with a double-base substitution event, an omikli event, or as a kataegis event.
